# Spatial biomarkers of response to neoadjuvant therapy in muscle-invasive bladder cancer: the DUTRENEO trial

**DOI:** 10.1101/2025.02.07.25321742

**Authors:** Enrique Grande, Mustafa Sibai, Elena Andrada, Daniela Grases, Oscar Reig, Marc Escobosa, Ainara Azueta, Daniel Castellano, Javier Puente, Jaime Martínez de Villarreal, Albert Font, Teresa Alonso-Gordoa, Raquel Benítez, Ane Moreno-Oya, Mario Álvarez-Maestro, Javier Burgos, M. Angel Climent, Mario Domínguez, Patricia Galván, Isabel Galante, Juan F. García, Elena Perez, Xavier García del Muro, Félix Guerrero-Ramos, Miriam Marqués, Pablo Maroto, Jesús M. Paramio, Alvaro Pinto, Aleix Prat, Núria Malats, Ignacio Durán, Eduard Porta-Pardo, Francisco X. Real

**Author notes:** **Correspondence:** Francisco X. Real, Spanish National Cancer Research Centre-CNIO, Melchor Fernandez Almagro 3, 28029-Madrid, Spain; email -, Eduard Porta, Institut Josep Carreras, Camí de les Escoles, s/n, 08916-Badalona, Spain, Enrique Grande, MD Anderson Cancer Center Madrid, Arturo Soria 270, 28033-Madrid, Spain. Equivalent contributions (Enrique Grande, Mustafa Sibai). Equivalent contributions (Elena Andrada, Daniela Grases).

## Abstract

Many studies have reported biomarkers predictive of response to immune checkpoint inhibitors (ICI) based on retrospective analyses. However, few clinical trials have tested their value prospectively. The DUTRENEO trial (EudraCT: 2017-002246-6) investigated whether an 18-gene Tumour Inflammation Signature (TIS) that can robustly identify patients who respond to ICI in multiple tumour types could stratify patients with localized muscle-invasive bladder cancer (MIBC) to receive neoadjuvant ICI (durvalumab+tremelimumab) or standard cisplatin-based neoadjuvant chemotherapy (NAC). Patients with TIS-high tumors were randomized to ICI or NAC, while patients with TIS-low tumors received NAC. A total of 73 patients were treated. Pathological complete response (pCR) rates were 38.5% (TIS-High, ICI), 30% (TIS-High, NAC), and 55% (TIS-Low, NAC) (p = 0.349), indicating that - as applied - the TIS score did not significantly enrich in responders to ICI. Post-hoc analysis showed that higher TIS thresholds improved prediction of response to ICI but excluded many responders. Multi-omics analyses of pre-treatment samples, including whole-exome sequencing, bulk RNA sequencing, and spatial transcriptomics (Visium, Xenium), revealed that bulk RNA response signatures originated mainly from cancer cells. Spatial transcriptomics showed that ICI response was associated with proximity between cancer cells and adaptive immune cells, while resistance to NAC was linked to phenotypic plasticity of cancer cells despite low genetic diversity. The unique design of the DUTRENEO trial underscores the challenges of translating retrospective biomarkers into clinical practice and highlights the importance of spatial features in understanding tumour-immune interactions. These findings suggest that integrating spatial and multi-modal biomarkers in well-designed clinical trials could improve stratification and response prediction to select neoadjuvant therapy.

## Introduction

More than 570,000 new cases of bladder cancer are diagnosed every year worldwide, causing approximately 200.000 yearly deaths^1^. While most of them are non-muscle invasive tumours (75%), a subset of patients (25%) present with muscle-invasive bladder cancer (MIBC), an aggressive disease. Radical cystectomy remains the most common therapeutic approach, although cisplatin-based neoadjuvant chemotherapy (NAC) provides an absolute 5-8% increase in 5-year overall survival (OS) and a 16% relative reduction in the risk of cancer-related death^2–6^. Recently, the NIAGARA study has shown that adding the anti-PDL1 immune checkpoint inhibitor (ICI) durvalumab to the standard regimen of cisplatin and gemcitabine as neoadjuvant therapy, followed by durvalumab for one year after cystectomy, increases the OS of patients. This regimen is expected to be incorporated soon into international guidelines for the management of these tumors^7^. Despite this evidence, real-world data reveal that there is a significant underuse of NAC, possibly related to concerns about treatment-related toxicity, the relatively modest absolute survival benefit, and the fact that 40-50% of patients are medically ineligible to receive cisplatin-based NAC^8^.

Small single-arm phase II trials using ICI doublets have revealed pathological complete response (pCR) rates in the range of 37-46%^9–12^. Retrospective analyses in several tumour types, including urothelial bladder cancer, have identified gene expression profiles associated with T cell infiltration, high interferon gamma (IFN-g), and low TGFβ signaling^13–15^ as predictive biomarkers of response to ICI. Particularly, a tumour inflammation signature (TIS) score based on 18 genes related to IFN-g signaling has been reported^15^. In these studies, the TIS score is one of the most robust biomarkers predictive of response to ICI, and its ability to discriminate is preserved across tumour types. The TIS has also been associated at the PanCancer level with prognosis and has been proposed as an indirect measure of an inflamed tumour phenotype that might guide immunotherapy across cancer types^16,17^.

A major problem in translational research is that few of the promising markers identified in retrospective studies, including the TIS, are applied prospectively to demonstrate clinical utility to stratify and optimize patient treatment. To our knowledge, the DOMINI trial in patients with stage III melanoma is the only study that randomized patients to allocate treatment according to a signature predictive of response to ICI^18^. In that trial, patients whose tumours had a high IFN-γ score showed a higher response rate than those with a low score, suggesting that the latter required additional or different therapies^18^. However, this study did not include a control arm of standard therapy.

Here, we present the results of the DUTRENEO trial which was uniquely designed to prospectively test the usefulness of stratifying patients with MIBC based on the TIS to select patients who could benefit more from neoadjuvant ICI, and to compare this arm to conventional NAC. Patients with a high TIS score tumour (immune-hot) were randomized to receive an ICI combination consisting of durvalumab and tremelimumab (ICI) or standard cisplatin-based NAC, according to the physician’s choice. To acquire a better understanding of the value of the signature, we also included patients whose tumours had a low TIS score (immune-cold) and who were treated with standard cisplatin-based NAC. The TIS, with the thresholds that we used, did not allow selecting patients with a higher response rate to ICI. To better understand the relationship between the molecular and cellular landscape of MIBC and the response to neoadjuvant treatment, we used multiple omics platforms (whole-exome sequencing - WES-, bulk RNA-Seq, and spatial transcriptomics). Spatial transcriptomics revealed that the cellular neighbourhood of cancer cells contains features associated with the response to ICI. In contrast, the phenotypic heterogeneity of cancer cells associates with the response to NAC. Overall, we highlight the challenges of translating retrospective biomarkers into the clinical setting and discover novel candidate mechanisms explaining the sensitivity or resistance of MIBC to different neoadjuvant therapies.

## Results

### Study design, patient characteristics, and safety

The DUTRENEO trial was a multicenter, open-label phase 2 study conducted in 11 Spanish tertiary care hospitals (**Figure 1a**). Patients with cT2-4N0-1 stage bladder cancer who had residual disease after transurethral resection of a bladder tumour (TURBT), were eligible for cisplatin therapy, had a planned cystectomy as part of their treatment plan, were ECOG 0-1 performance status, and had tumour tissue available for TIS quantification at baseline were recruited. One-hundred-one patients were screened for study eligibility. Of them, 28 resulted in screening failures (**Extended Data Figure S1a, Methods**). Upon establishment of the TIS score, patients whose tumours were TIS-high (i.e., hot) were randomized to receive ICI or standard cisplatin-based NAC. In contrast, patients whose tumours were TIS-low (i.e., cold) were assigned to receive standard NAC (**Methods**). Overall, 73 patients were eligible and received treatment, as follows: 21 patients with cold tumours received cisplatin-based NAC according to the investigator’s choice. Fifty-two patients had hot tumours. The first 6 patients were treated with durvalumab plus tremelimumab under a safety run-in period for at least two cycles, and the remaining 46 patients were randomized to ICI (n=24) or NAC (n=22) (**Extended Data Figure S1a**). Cystectomy, post-surgery follow-up, and therapy were done according to standard practice. Clinical and pathological baseline characteristics of patients in the three arms were not significantly different (**Table 1**). Adverse effects and toxicities were in line with those previously described for NAC and ICI (**Extended Data Table 1**).

**Figure 1.**
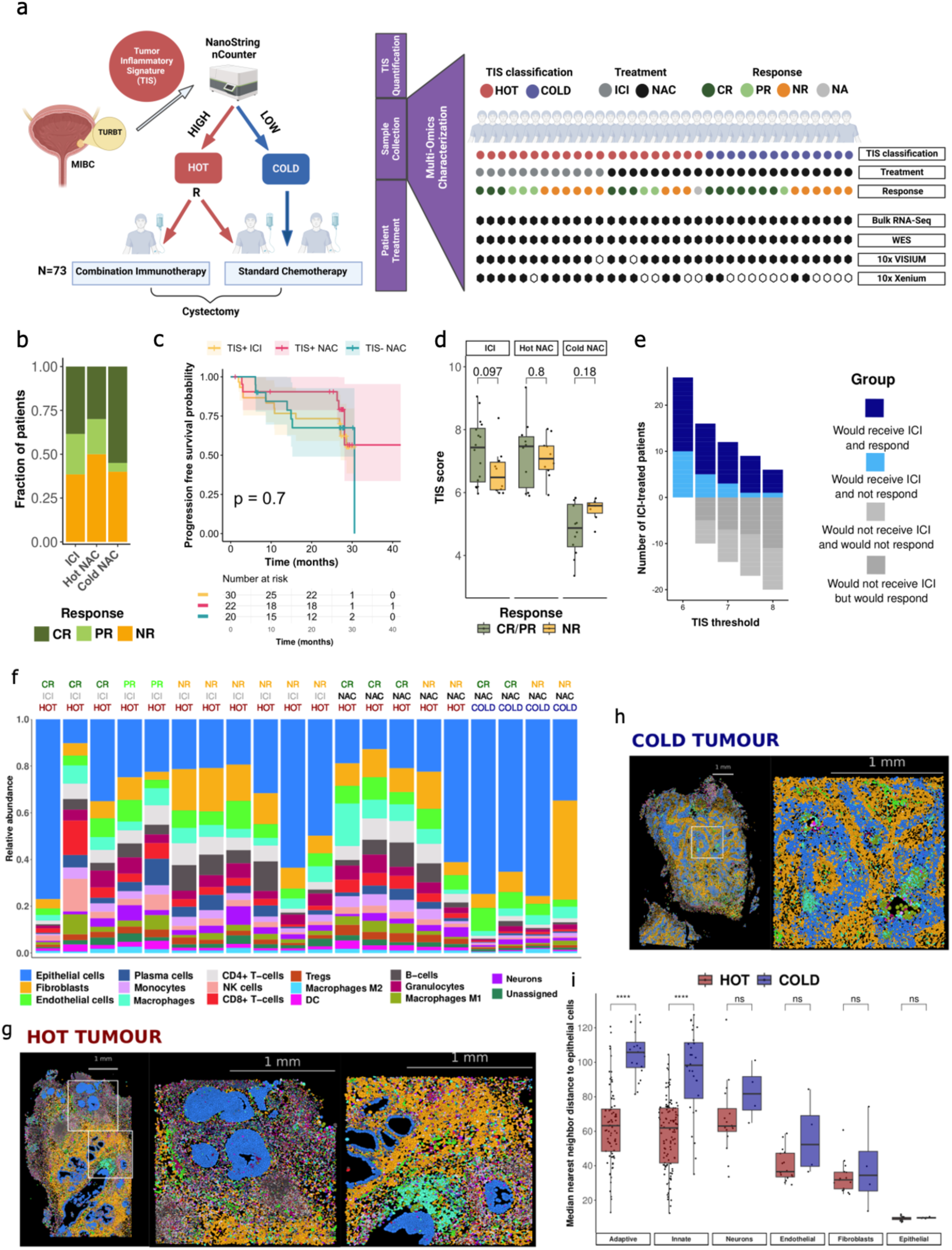
Study design, outcome, and multiomics analysis. **a)** DUTRENEO study design. Patients’ tumours were classified as immune-hot (hot, for simplicity) or immune-cold (cold, for simplicity) based on the score of the TIS signature. Patients with hot tumours were randomized to receive ICI or NAC whereas patients with cold tumours received standard NAC. Multiomics analyses of pre-treatment tumour samples were performed using WES (n=44), bulk RNA sequencing (n=57), as well as spatial transcriptomics (n=35). For spatial transcriptomics, data were generated from samples representative of the three study arms using Visium (n=33) and Xenium (n=20) platforms, 18 of which come from the same patient. **b)** Barplot showing the fraction of patients who had complete (CR), partial (PR) or no (NR) pathological response in each of the treatment arms. **c)** Kaplan-Meier plot showing progression-free survival for patients included in the three treatment arms. **d)** Boxplot showing that the TIS threshold does not discriminate well responders from non-responders in any of the treatment arms. **e)** Barplot showing the retrospective analysis of DUTRENEO outcomes. The y-axis shows the fraction of patients that would have been included (positive values) or excluded (negative values) to receive neoadjuvant immunotherapy at different TIS thresholds (x-axis). Bars are colored depending on the outcome of the patient. **f)** Relative cell type abundance in the samples analyzed with the Xenium platform. **g, h)** Overlay of the major cell types in the whole sample analyzed and in selected regions of exemplary hot (**g**) and cold tumours (**h**). **i)** Boxplot showing the median nearest neighbour distance of the different cell types to epithelial cells (calculated within a 200 μm radius) in samples analyzed with the Xenium platform, split by TIS category. Median distances of individual cell types pertaining to the adaptive and innate immune system were grouped under “adaptive” and “innate”, respectively.

**Table 1.**
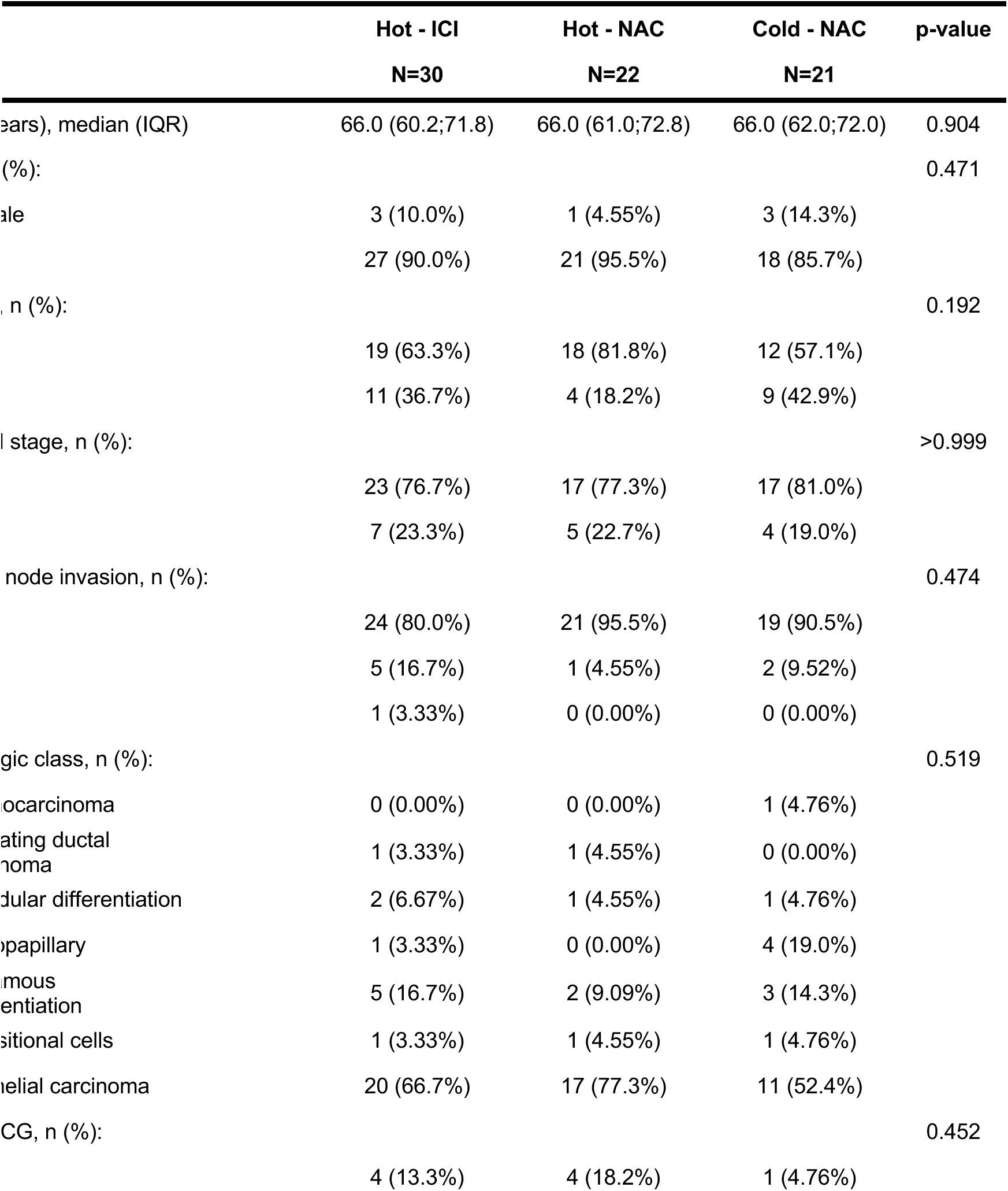
Baseline characteristics of patients recruited to the DUTRENEO study.

### Efficacy outcomes

Among patients with hot tumours, a pathological complete response (pCR) was observed in 10/26 patients treated with ICI (39%) and in 6/20 patients treated with NAC (30%). Among patients with cold tumours, 11/20 had a pCR (55%) (**Table 2**, **Figure 1b**). There was no statistically significant difference in the primary endpoint between patients with hot tumours treated with ICI and those treated with NAC (Chi-squared p > 0.8) nor between patients treated with NAC who had hot or cold tumours (Chi-squared; p> 0.5). Downstaging was achieved in 10/20 (50%) and 12/20 (60%) patients with hot or cold tumours, respectively, and in 16/26 (62%) of patients treated with ICI. There were no significant differences in progression-free survival (p = 0.7, **Figure 1c**) and overall survival (p > 0.6, **Extended Data Figure S2a**) across treatment groups. We did not find significant differences in the TIS score of responders vs. non-responders in any of the treatment arms, although there was a tendency for higher scores in responders from the ICI arm (p > 0.097) (**Figure 1d**). Finally, the pCR rate was similar to that observed in other studies assessing ICI in an unselected population of MIBC, suggesting that the TIS, as used in this trial, did not enrich the treated population with responders. A retrospective analysis of the outcome data of DUTRENEO shows that higher TIS thresholds would have increased the fraction of responding patients at the expense of excluding most patients who responded to the neoadjuvant immunotherapy (**Figure 1e**).

**Table 2.** Pathological response and downstaging rates^a,b^.

|  | Hot - ICI<br>N=26 | Hot - NAC<br>N=20 | Cold - NAC<br>N=20 | p-value |
| --- | --- | --- | --- | --- |
| Pathological response, n (%): |  |  |  | 0.349 |
| Complete response | 10 (38.5%) | 6 (30.0%) | 11 (55.0%) |  |
| Partial response | 6 (23.1%) | 4 (20.0%) | 1 (5.00%) |  |
| No response | 10 (38.5%) | 10 (50.0%) | 8 (40.0%) |  |
| Downstaging, n (%): |  |  |  | 0.710 |
|  | 16 (61.5%) | 10 (50.0%) | 12 (60.0%) |  |

<sup>a</sup> Cystectomy was not performed in 4 patients from the hot ICI group (two patients progressed while receiving treatment, one patient refused cystectomy, one developed myocarditis) and 1 patient from the cold group who was incorrectly enrolled in this group. One patient from the hot chemotherapy group withdrew consent to use information on cystectomy outcome; a second patient developed grade 3 rash leading to a delay in cystectomy beyond protocol rules.
<sup>b</sup> Downstaging was defined as any improvement in T or N stage from the time of randomization.

### A comprehensive multi-omics characterization of the DUTRENEO cohort

We used multiple molecular platforms to characterize the pre-treatment samples and improve the prediction of response to therapy (**Figure 1a**). Formalin-fixed paraffin-embedded (FFPE) tissues obtained during the TURBT were used for WES (n = 44), bulk RNA sequencing (n = 57), and spatial transcriptomics with both Visium (n = 33) and Xenium (n = 20) to achieve whole-transcriptome coverage and single-cell resolution (**Figure 1a**). The Visium data included 99,028 spots across all samples (median, 3147 per sample) and an average number of 4,097 unique genes were detected per spot. The Xenium data, on the other hand, included 377 genes analyzed in 5,399,355 cells (median, 224,001 cells per sample), with a median number of 48 transcripts and 28 genes per cell.

To determine whether the patient cohort is representative of primary untreated MIBC, we considered the frequency of cancer driver mutations and the molecular subtypes. The frequency of driver mutations in the main bladder cancer genes (e.g., *TP53*, *FGFR3*, *PIK3CA*, *KMT2D*) is similar to that described in larger cohorts^19^ (**Extended Data Figure S1b**). Consensus molecular subtype distribution is also similar to that reported previously^20^, except for a predominance of luminal subtypes likely reflecting the use of TURBT samples: LumP (n=20), LumNS (n= 9), LumU (n=7), Stromal-rich (n=9), and Ba/Sq (n=12) (**Extended Data Figure S1c**). As expected, *FGFR3* mutations were enriched among LumP tumours. Subtype distribution was not associated with treatment received or response to therapy (**Extended Data Figure S1d-f**).

Comparison of hot and cold tumours reveals that mutations in *TP53*, *RB1*, and *ARID1A* are more common in hot tumours, whereas *FGFR3* and *STAG2* mutations are more common in cold tumours^19^ (**Extended Data Figure S1b**). RNA-Seq analysis of differentially expressed genes (DEG) in hot, compared to cold tumours is consistent with the stratification criteria: in hot tumours, overexpressed transcripts corresponded to immune-related genes (e.g., *IDO1*, *IFNG*) and pathway analysis confirms enrichment of immune pathways (e.g., T cell receptor signaling, NK-mediated cytotoxicity, PD-1 signaling) (**Extended Data Figure S3a**). Among the genes significantly overexpressed in cold tumours are *MYCN*, *MYCL*, and *FGFR3*. This gene-set was enriched in pathways involved in DNA repair (e.g., homologous recombination, nucleotide and base excision repair) and RNA and chromatin biology. Hot tumours showed an over-representation of Stromal-rich and Basal-squamous subtypes whereas cold tumours were enriched in the luminal subtypes, especially the LumP subtype (**Extended Data Figure S1d**).

To acquire more resolution on the molecular features of the tumour and its microenvironment, we used Xenium to profile the expression of a panel of genes designed to annotate cell types from multiple tissues (**Methods**). We found that the cellular composition of tumours was very heterogeneous (**Figure 1f-h**). Among hot tumours, there were no major differences in both treatment arms (**Extended Data Figure S4a**). For example, the fraction of epithelial cells ranged between 9% and 72% and that of fibroblasts between 3% and 33%. The cell composition correlated with the TIS score, as hot tumours had on average a higher fraction of immune cells than cold tumours: B cells (3.7% vs 0.9%, p = 0.015), CD4 T cells (4.6% vs 1.4%, p = 0.004), Tregs (2.1% vs 0.7%, p = 0.001), and CD8 T cells (4% vs 0.7%, p = 0.0004) (**Extended Data Figure S4b**). Moreover, the median distance between epithelial and the nearest neighbouring immune cells was significantly shorter in hot than in cold tumours for both the adaptive (mean, 63 vs 104 µm, p < 0.001) and innate (mean, 58 vs 94 µm, p < 0.001) immune groups (**Figure 1i**). This shows that cold tumours not only have fewer immune cells overall (**Figure 1f**) but also that they are located further away from cancer cells (**Figure 1i**). Remarkably, the distance between cancer and non-immune cells of the TME, including fibroblasts, neurons or endothelial cells, was not significantly different (**Figure 1i**). Overall, the TIS signature discriminates between hot vs. cold tumours and captures selected features of the TME.

### Spatial transcriptomics reveals new insights into chemotherapy sensitivity

To identify transcriptomic features associated with response to therapy and overcome the limitations of the TIS classification, we leveraged the bulk RNA-seq and ST data. Comparison of the transcriptome of patients treated with ICI reveals 858 over-expressed and 3432 under-expressed in responders (p-adj < 0.05, log2FC > 1 or < −1). Among the genes with significantly higher expression in responders are *AQP3*, *KRT4*, *PRC1*, *CXCL10*, *IFNG*, *MCM2*, and others involved in T cell function (**Figure 2a**). Pathways significantly enriched include those related to cell cycle (e.g., p53 signaling, E2F targets, *MYC* targets, and G2/M), oxidative phosphorylation, DNA repair, and signaling by FGFR and MAPK (**Extended Data Figure S3b**). Among patients treated with NAC (both arms), 607 DEGs are differentially expressed in responders vs. non-responders (64 over-expressed and 543 under-expressed). Among the former are *TAC3*, *SIGLEC6*, *CHST9*, *FGFR3*, *MYCL*, or *GATA2* (**Figure 2a**). This gene-set is significantly enriched in pathways related to EGFR and FGFR signaling, RNA biology, endoplasmic reticulum, oncogene-induced senescence, and cholesterol biosynthesis (**Extended Data Figure S3c**).

**Figure 2.**
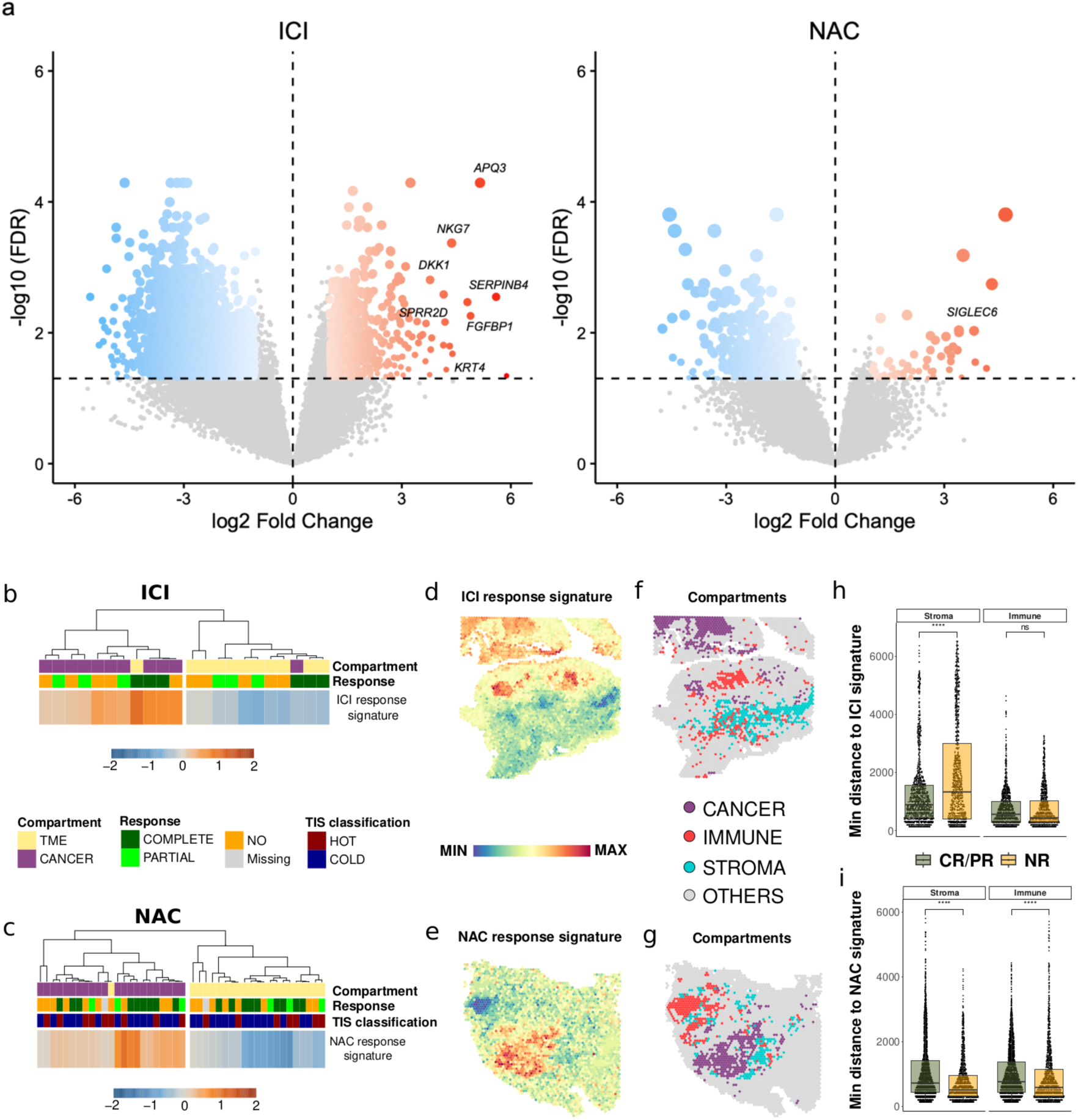
Spatial transcriptomics reveals new insights from bulk-derived expression signatures. **a)** Volcano plots of the bulk-derived differentially expressed genes between responders and non-responders to ICI (left) or NAC (right). **b,c)** Hierarchical clustering of the samples analyzed using Visium in patients receiving ICI (**b**) or NAC (**c**) showing the contribution of the cancer vs. TME compartments (as calculated by ESTIMATE) to the quantified module score of bulk-derived upregulated genes (i.e., response signatures) in responders to ICI or NAC. **d,e)** Overlay of the response signatures on two exemplary samples from patients who responded to ICI (**d**) or NAC (**e**). **f,g**) Overlay of the cancer, immune, and stromal compartments on the same samples shown in (**d** and (**e**), respectively. Spots were given their labels by taking the 90th percentile of corresponding continuous scores generated by ESTIMATE. **h,i)** Boxplots showing the minimum distance of all stromal or immune spots to the respective response signatures in the ICI (**h**) or NAC (**i**) cohorts.

Next, we investigated the distribution of the bulk transcriptomics-derived signatures of genes overexpressed in responders to ICI or NAC (compared to non-responders; log2FC > 1 and p-adj < 0.05) at the spatial level using Visium data. We used ESTIMATE^21^ to define the cancer and TME compartments and assess their contribution to the corresponding upregulated genes (i.e. response signatures). These signatures originated from the cancer compartment in both responders and non-responders to both ICI and NAC (**Figure 2b,c**). To establish the spatial relationship with the TME, we calculated the minimum distance of all stromal or immune spots to the response signature spots (**Figure 2d-g**). In patients who received ICI, the minimum distance of stromal spots to the ICI response signature spots was significantly shorter in responders compared to non-responders (**Figure 2h**). In this group of patients, the minimum distance of immune spots to the ICI response signature spots was not predictive of response to ICI (**Figure 2h**). In contrast, in patients who received NAC the minimum distance of either stromal or immune spots to the chemotherapy response signature spots was significantly greater in responders than in non-responders (p < 0.0001, **Figure 2i**). These findings support the existence of spatially defined cancer cell communities with distinct transcriptional programmes associated with response to ICI or NAC.

### Intratumoural heterogeneity correlates with resistance to NAC

Intratumoural heterogeneity (ITH) is a major determinant of therapeutic resistance^22,23^. ITH can originate from genetic or non-genetic mechanisms. While the former refers to the genomic differences between clones, the latter refers to phenotypic variation in the absence of major differences at the DNA level (i.e., plasticity). The two types of ITH are not mutually exclusive.

The Visium data allowed us to explore the association of both types of ITH with therapeutic response. To estimate the degree of genetic ITH, we used InferCNV^24^, which identifies the presence of different clones in each tumour sample, as well as their genetic differences, calculated as the fraction of the genome in which two clones differ in their CNV state. We found an average of 6 clones per tumour (range 1-18), (**Extended Data Figure S5a**). Based on the distribution of genetic differences, we defined two groups of pairs of clones. The first group, which we designate as genetically similar, differs in their CNV state by less than 10% of the genome. These represent 70% and 51% of all pairs in cold and hot tumours, respectively (**Figure 3a**). The second group, genetically distant, differs in the CNV state in more than 10% of their genome, and are 30% and 49% of all pairs in cold and hot tumours, respectively. Finally, we used these two measures (number of clones per sample and maximum genetic difference between a pair of clones of the same sample) as a proxy for genetic ITH in each sample and tested their association with response to therapy. The number of clones per tumour was similar in responders and non-responders in both groups (**Extended Data Figure S5b**). However, patients who responded to NAC tended to have pairs of clones that are more genetically distinct (p=0.007, **Extended Data Figure S5c**). This was not the case in patients who received ICI (p=0.33, **Extended Data Figure S5c**). This suggests that genetic ITH, measured as CNV, could be predictive of response to NAC, but not to immunotherapy.

**Figure 3.**
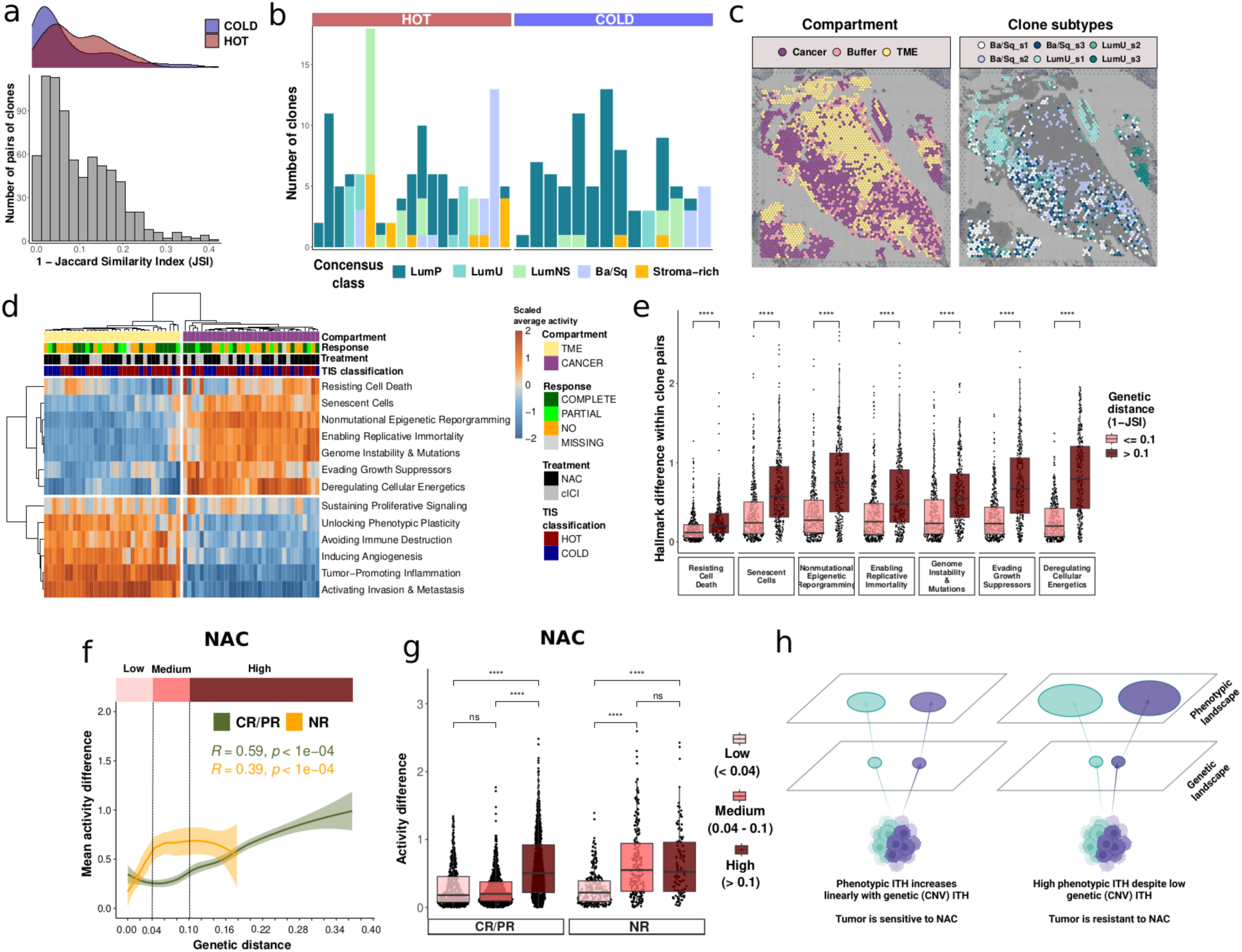
Intratumoural heterogeneity is associated with resistance to NAC. **a)** Distribution of the genetic distances between pairs of predicted clones. Density plot showing the distribution based on the TIS classification (top) and histogram showing Jaccard similarity index (bottom) (n=33). **b)** Number of predicted clones in each sample, coloured by the assigned consensus molecular subtype. **c)** Distribution of the tumour and TME compartments in the sample of a patient who received NAC (left) and distribution of the predicted clones according to the assigned molecular subtype (right). **d)** 2D hierarchically clustered heatmap showing the scaled average Cancer Hallmark scores (rows) within an ESTIMATE cluster across the spots in each sample (columns) (n=33). **e)** Neoplastic Hallmark activity difference within pairs of predicted clones stratified according to their genetic distance based on Jaccard similarity index (1-JSI) (n=32). **f)** Scatter plot of the genetic distance and the activity difference within clone pairs across six (all, except for “Deregulating cellular energetics”) neoplastic Hallmarks in the samples of patients who received NAC (hot and cold combined), split by treatment response. Three windows of genetic distance were highlighted to reflect the observed patterns (dashed vertical lines at 0.04 and 0.1). Pairs of clones with genetic distance <0.4 were classified as “low”, those between 0.04 and 0.1 as “medium”, and those >0.1 as “high”, and were all coloured in bright, medium, or dark red, respectively, with horizontal bars at the top. **g)** Activity difference of the 6 neoplastic hallmarks within pairs of clones, split by the genetic distance windows defined in panel f, further split by response to ICI. **h)** Graphic summary findings of panels **f-g**, where non-responders to NAC display high phenotypic ITH despite low genetic ITH (based on CNV calculations), whereas responders show a linear relationship between phenotypic and genetic ITH. *ns: p > 0.05, *: p <= 0.05, **: p <= 0.01, ***: p <= 0.001, ****: p <= 0.0001*.

To assess the contribution of non-genetic ITH to therapeutic sensitivity, we first considered whether tumors belonging to different molecular subtypes can be identified within a single tumour. We assigned a molecular subtype to each clone in each sample using a mini-bulk RNA-Seq from all spots corresponding to the clone. Overall, in 45% of the samples (15/33) all clones belong to the same molecular subtype; in 42% of cold (6/14) and 63% of hot (12/19) tumours, at least one clone of a different subtype was identified (**Figure 3b**). Clones belonging to different molecular subtypes are often segregated in space (**Figure 3c**). There was no association between the existence of clones belonging to multiple molecular subtypes in a sample and response to therapy in any of the treatment groups: ICI (p = 0.24), NAC (p = 0.36). Similarly, using the activity of the Cancer Hallmarks^25^ as a phenotype proxy for each clone (**Figure 3d**), we did not find significant differences between responders and non-responders in any of the trial’s arms (**Extended Data Figure S5d**). Overall, our results showed that phenotypic ITH, on its own, does not associate with response to therapy.

Finally, we explored the interplay between genetic and phenotypic ITH and its association with treatment response. Overall, there is a positive association between genetic and phenotypic heterogeneity: pairs of clones with high genetic differences differ more in the activity of Cancer Hallmarks, and vice versa (**Figure 3e**). Next, we explored whether this association varied according to the response to therapy. In patients who received ICI, we found no differences (**Extended Data Figure S5e**). However, the tumours of patients who received NAC and did not respond contained cancer clones that displayed high phenotypic diversity despite minimal genetic differences (**Figure 3f,g**). This was not the case in tumours of NAC responders, where the relationship between genetic and phenotypic ITH was more gradual, as observed in all ICI patients. This suggests that tumours with high plasticity and low genetic diversity (measured as CNV differences) are more likely to be intrinsically resistant to chemotherapy (**Figure 3h**).

### Immune checkpoint gene expression and response to ICI

Some studies have suggested that the expression of the checkpoint genes/proteins could be used as a biomarker to predict the success of checkpoint blockade^26^. We explored to what extent this could be used to predict the outcome of DUTRENEO patients treated with neoadjuvant therapy. Protein levels of PDL1, assessed using IHC, were not associated with the main clinical and pathological features (e.g., T stage, grade or lymph node involvement) or response to therapy (not shown). Using bulk RNA-seq data, tumours of responders have significantly higher levels of *PDL1* (log2 fold-change > 1.8, p-value = 0.001) and *CTLA4* (log2 fold-change > 2, p-value = 0.002) than non-responders.

We next explored the distribution of *PDL1* and *CTLA4* at the single cell level using the Xenium data. We first considered which cell types contribute the most to the overall expression of *PDL1* and *CTLA4*. In the case of *PDL1*, we observed two patterns, with 4/11 patients having >50% of transcripts coming from cancer cells and 6/11 having <25% (**Figure 4a**). This bimodal distribution did not associate with sensitivity to ICI (**Extended Data Figure S6a**). As for *CTLA4*, Tregs are the predominant source in the 11 samples analyzed, consistently contributing the highest proportions, ranging from 13-34% (**Extended Data Figure S6b**). Other CD4 T cells and epithelial cells also accounted for 9-37% and 2-52%, respectively. While the input of CD8 T cells was relatively lower overall, ranging from 2-30%, their expression of *CTLA4* was associated with response to ICI therapy (p = 0.004, **Extended Data Figure S6c**). Other cell types had negligible contributions, emphasizing the cell-type specificity of *CTLA4* expression and the distinct role of CD8 T cells in therapy response.

**Figure 4.**
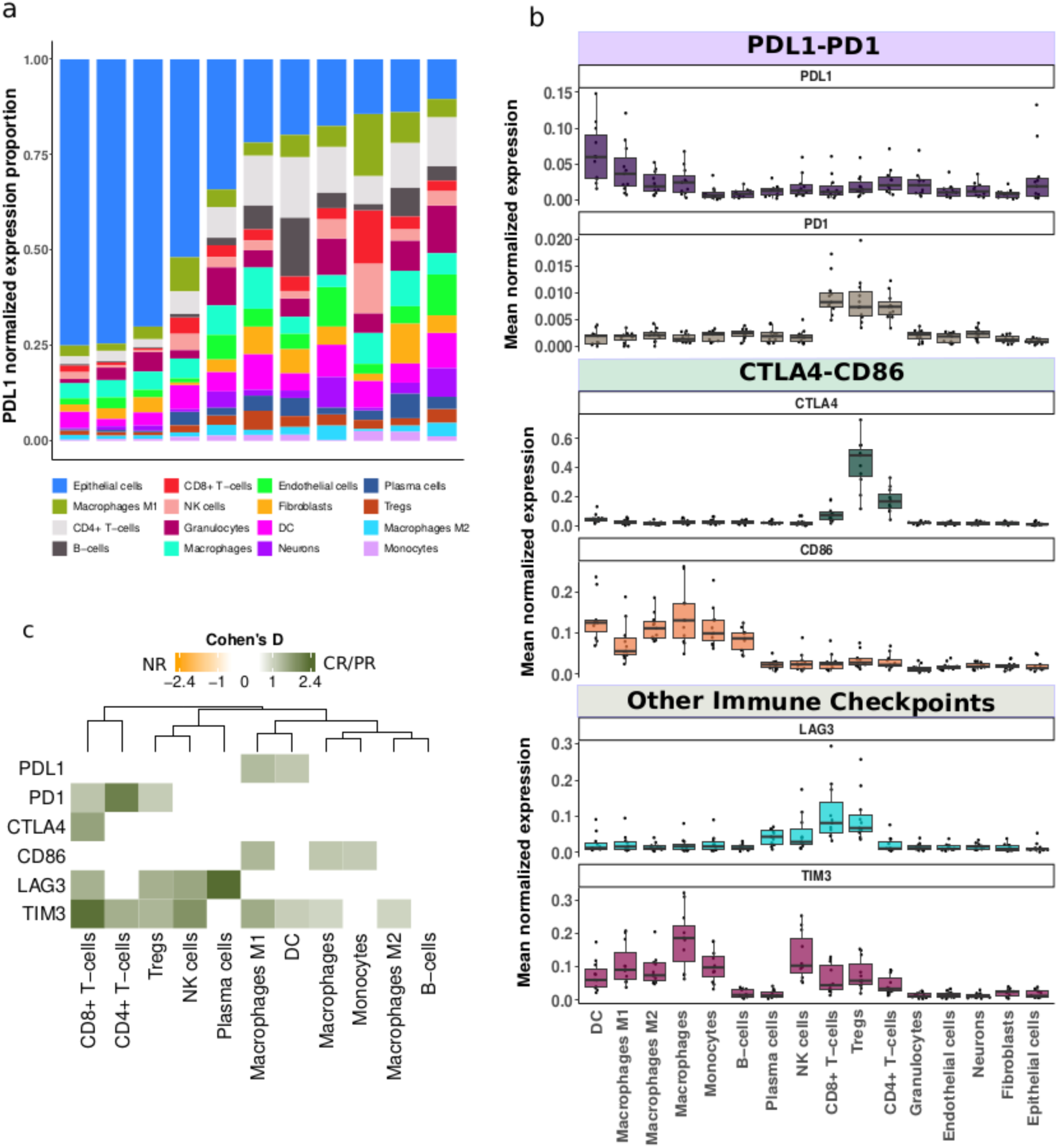
Gene expression landscape of immune checkpoint genes and their partners in ICI-treated patients. **a)** Cell type distribution of *PDL1*-expressing cells (normalized expression). **b)** Distribution of the mean normalized expression of *PDL1*, *PD1*, *CTLA4*, *CD86*, *LAG3*, and *TIM3* across samples for each cell type. **c)** Heatmaps showing Cohen’s D of the mean normalized expression of *PDL1*, *PD1*, *CTLA4*, *CD86*, *LAG3,* and *TIM3* in their relevant cell types (based on data from panel b) between responders and non-responders. Higher Cohen’s D (dark green) indicates a higher mean expression in the relevant cell type in responders, whereas a higher value towards the orange color indicates the opposite.

To further explore the potential association of checkpoint gene expression and ICI outcomes at the single cell level, we examined the cell types having higher mean expression of *PDL1* and *CTLA4*, their interaction partners *PD1* and *CD86*, and *LAG3* and *TIM3*, two other checkpoint genes. Regarding *PDL1*, the cell type with the highest mean expression were dendritic cells followed by M1 macrophages (**Figure 4b**), whereas its binding partner *PD1* was highly expressed in multiple T cell types. This was also the case of *CTLA4*, with high expression in CD8, CD4 and T regulatory cells, while the expression of *CD86*, its binding partner, was highest in antigen-presenting cells: M1 and M2 macrophages, monocytes, DCs, and B cells (**Figure 4b**). Regarding the other checkpoints, the mean expression of *LAG3* was highest in CD8 T cells, Tregs, plasma, and NK cells, whereas that of *TIM3* was high in all immune cells except for B cells, plasma cells, and granulocytes (**Figure 4b**). Finally, we used Cohen’s D to test whether responders and non-responders to ICI differ in the expression of these genes in the cell types where they are highly detected. Overall, the mean expression of all six genes in their relevant cell types was higher in patients who responded to ICI (**Figure 4c**). For example, the mean expression of *PD1* on CD4 T cells and that of *CTLA4* on CD8 T cells was higher in responders (p = 0.008 and p = 0.03, respectively, **Figure 4c**). Similarly, the mean expression of *TIM3* on CD8 T cells and that of *LAG3* on plasma and NK cells was higher in responders (p = 0.03, p = 0.008, p = 0.01, respectively, **Figure 4c**). Overall, these results suggest that bulk expression levels of checkpoint genes are associated with sensitivity to ICI and their expression in biologically relevant cell types further refines response prediction.

### Cellular neighbourhoods predict ICI response

We explored the location of the cell types relevant to ICI response within the tumour. First, we calculated the median distance between epithelial cells and their nearest neighbouring cell types within a 200 μm radius of each epithelial cell. In responder patients, epithelial cells tended to be significantly closer to immune cells of the adaptive (mean, 54 vs 75 μm, p < 0.001, **Extended Data Figure S7a**) and innate arms (mean, 53 vs 66 μm, p < 0.05, **Extended Data Figure S7a**) than in non-responders. Cell type composition analysis showed an association with outcome only for fibroblasts, which were more abundant in non-responders. Overall, this suggests that the location of specific immune cells within the tumour, rather than their overall abundance, predicts the success of ICI.

Next, we observed that the most frequent median nearest-neighbour distances to epithelial cells in the previously established 200 µm radius was approximately 25 µm in the ICI Xenium data (**Extended Data Figure S7c**). This suggests most cell-cell communication events occur around such distance. To further explore how cell-cell interactions influence the sensitivity to ICI, we used an unsupervised approach to identify local cellular neighbourhoods (**Figure 5a**). In short, we created a matrix in which we classified and quantified the cells within a 25 µm radius (the neighbourhood) of each cell (**Figure 5a**). Then, we used K-means clustering to identify cellular communities, which are groups of cells with a similar neighbourhood composition (**Figure 5b**, **Extended Data Figure S8**). Overall, we identified 68 communities in the 11 tumours (mean of 6 communities per sample). Next, we quantified the enrichment or depletion of each cell type within each community, allowing us to determine whether a community is “rich” (enriched), “neutral”, or “poor” (depleted) in any given cell type (**Figure 5b**). Finally, we grouped together all the “cell type-rich” communities within a sample (e.g., fibroblast-rich community), iterating this process for each individual cell type (**Figure 5c-g**).

**Figure 5.**
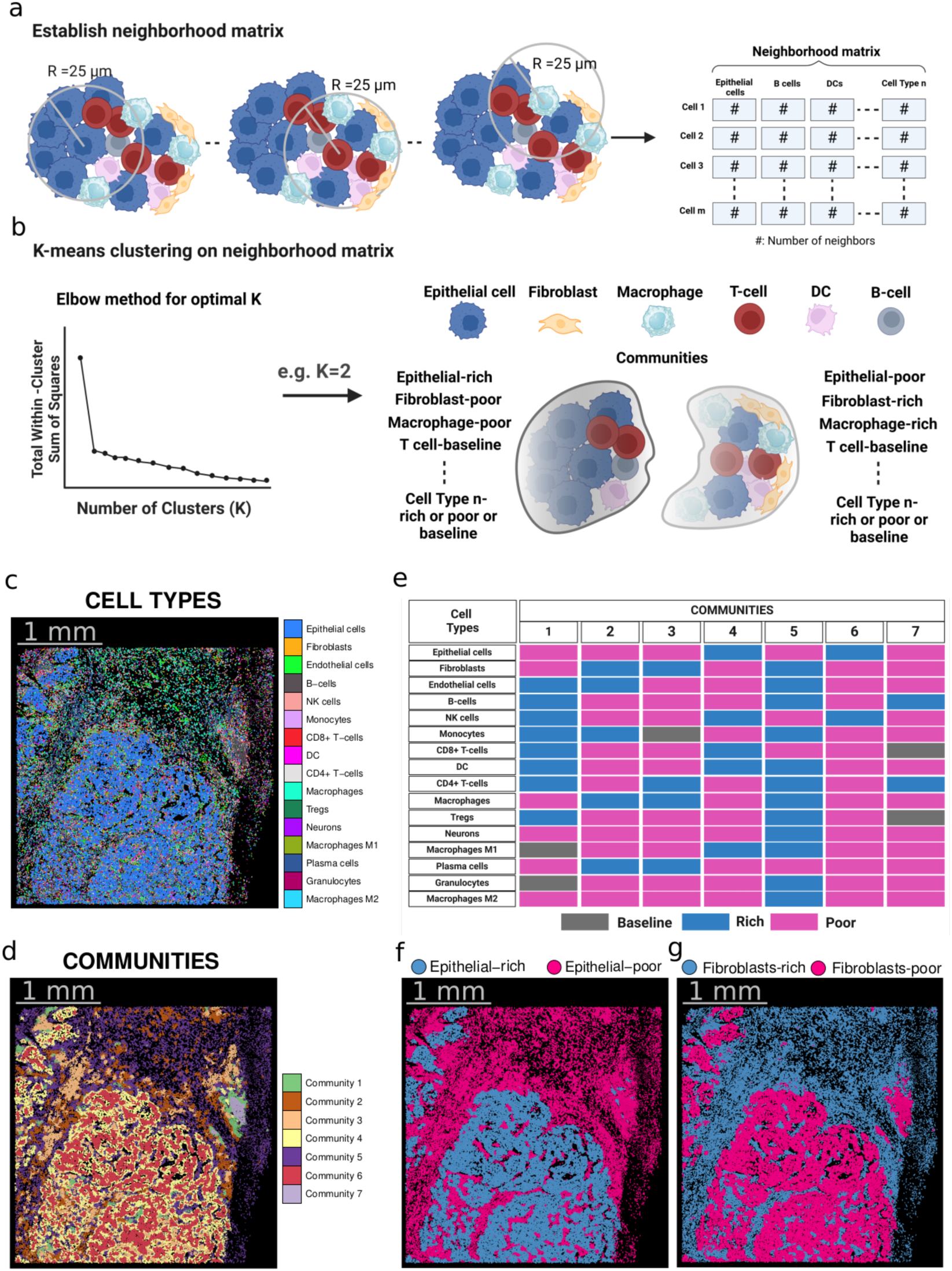
Neighbourhood analysis strategy. **a,b)** Scheme of the neighbourhood analysis for the identification of communities and their cellular composition. First, a neighbourhood radius of 25 µm is established around each cell, within which the number of cells of a given type are counted to form a neighbourhood matrix (**a**). Next, the elbow method is applied on the neighbourhood matrix for each sample to determine an optimum number of clusters (i.e., communities) that will be used for k-means clustering. The relative abundance of each cell type within each community is finally determined, revealing its level of abundance as “baseline” (neutral), “rich” (high abundance), or “poor” (low abundance) (**b**). **c-g)** Example of community identification steps from panel (**b**) on a section of a hot tumour, showing the overlaid cell types (**c**), the overlaid communities (**d**), the abundance of each cell type in each of the communities across the whole sample (**e**), the overlaid communities that are epithelial-rich or poor (**f**), and those that are fibroblast-rich or poor (**g**).

We then compared the relative abundance of individual cell types in each of the cell type-rich communities in responders and non-responders, aiming to identify compositional features associated with outcome. Overall, we found that dendritic cells, M1 macrophages, CD8 T cells, T regs, and NK cells were more prevalent in most communities of responding patients (**Figure 6a**). These associations were particularly strong for CD8, T regs, and NK cells in the epithelial-rich communities, and dendritic cells in monocyte-rich and endothelial-rich communities. On the other hand, non-responding patients had a higher relative abundance of fibroblasts in M1 and M2 macrophage-rich, and CD8-rich communities, suggesting that cancer-associated fibroblasts impact the efficacy of ICI.

**Figure 6.**
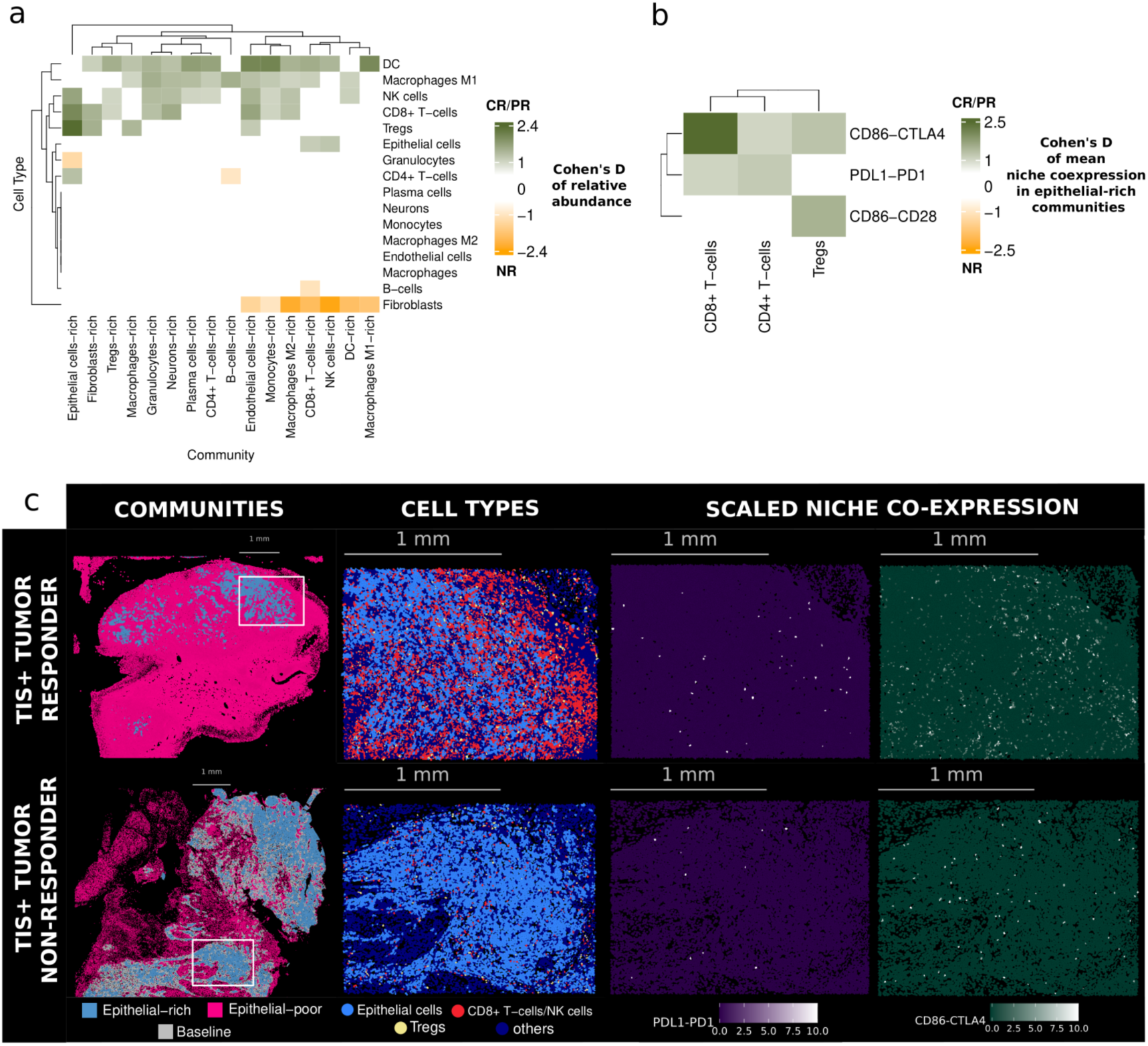
Cellular neighbourhoods predict ICI response. **a)** Heatmap showing Cohen’s D of the relative abundance of each cell type in each cell-rich community, comparing the two response groups among cases from the ICI Xenium cohort. Higher Cohen’s D (dark green color) indicates a higher relative abundance of a given cell type in each community in responders, whereas a higher value towards the orange color indicates the opposite. **b)** Heatmap showing Cohen’s D of the mean niche co-expression of the ligand-receptor (LR) pairs of interest. Higher Cohen’s D towards (dark green color) indicates a higher niche co-expression of an LR pair around a given T cell type in responders compared to non-responders, whereas a higher value towards the orange indicates the opposite. **c)** Selected findings in the tumours of one ICI responder (top) and one non-responder (bottom). From left to right: the segmented cells from epithelial-rich vs. epithelial-poor communities are shown, followed by the cell type composition of the insets from the previous community images, with a focus on epithelial cells, CD8 T cells/NK cells, and Tregs. The last two images refer to the scaled niche co-expression of LR pairs *CTLA4*-*CD86* and *PDL1*-*PD1* in the regions from the insets.

Given the differences in checkpoint expression across cell types, and the differences in cell type composition of the communities of cancer cells in responders and non-responders, we explored whether these two features had synergistic effects. This provides a means to dissect localized signaling mechanisms that might drive therapeutic efficacy. To this end, we quantified the niche co-expression of the most relevant ligand-receptor (LR) pairs (*PDL1-PD1*, *CTLA4-CD86*, and *CD86-CD28*) in epithelial-rich communities. We defined niche co-expression as the mean co-expression of a ligand and its receptor in the neighbouring three cells of a target cell, capturing the local signaling environment (**Extended Data Figure S9a**). The mean niche co-expression of *CD86* and *CD28* around *CD8* or non-regulatory CD4 T cells did not differ by response to ICI (**Figure 6b**). However, the mean niche co-expression of *CD86* and *CTLA4* around all T cell types tended to be higher in ICI responders compared to non-responders, especially around CD8 T cells (p < 0.01, **Figure 6b,c**). Additionally, the mean niche co-expression of *PDL1* and *PD1* around CD8 and non-regulatory CD4 T cells tended to be higher in responders compared to non-responders (**Figure 6b,c**). This association was more significant around the latter cell type (p < 0.05), even more so when only cases with a pCR were considered (p < 0.01, **Extended Data Figure S9b-c**). Together, these results highlight that the selective engagement of these ligands with their receptors on different types of T cells in cancer cell-rich areas may be crucial to determine the response to ICI.

### Checkpoint signaling is spatially confined to specific communities in responding patients

We hypothesized that the expression of immune checkpoints and their binding partners by a given cell type might vary depending on the cellular community in which the cell resides. Specifically, we aimed to determine whether these community-associated differences in immune signaling correlate with differential responses to neoadjuvant ICI. This could point to spatially-restricted immune signaling, or to cell-cell communication mechanisms that might be relevant to create a niche that can be targeted by ICI. To test this, we systematically compared the expression of immune checkpoints and their binding partners on each cell type within heterotypic cell-type rich and cell-type poor communities. As an illustrative example, we explored *PDL1* expression in M1 macrophages located within endothelial-rich and endothelial-poor communities (**Figure 7a,b**). In non-responding patients, M1 macrophages located in endothelial-rich communities contributed significantly more to *PDL1* expression than those in endothelial-poor communities (**Figure 7c**, adjusted p = 0.01), highlighting a spatially conditioned mechanism that may influence response.

**Figure 7.**
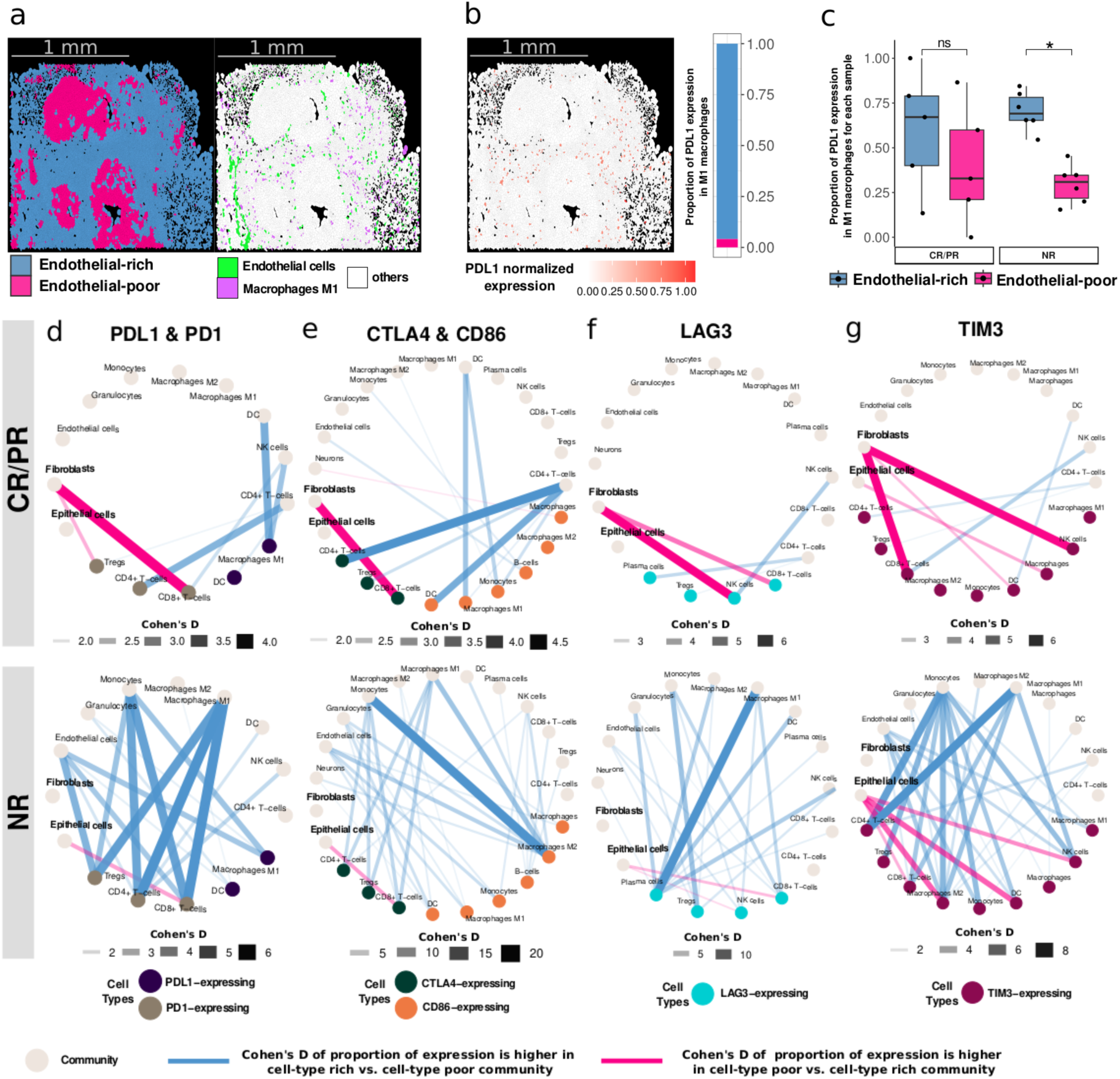
Spatially constrained checkpoint signaling associates with response to ICI. **a-c)** Examples of the distribution of PDL1 transcripts in M1 macrophages in endothelial-rich and endothelial-poor communities (i.e., the relative share of *PDL1* expression contributed by these two communities) on a section of a hot tumour. **a)** Overlay of the endothelial cell-rich and - poor communities on the left, followed by an overlay of endothelial cells (green), M1 macrophages (purple), and any other cell type (white) on the right. **b)** Overlay of the normalized expression of *PDL1*, followed by a barplot of the relative contribution of *PDL1* expression in M1 macrophages by endothelial-rich and endothelial-poor communities in that tissue section. **c)** The same contributions from (b) across the whole sections of each sample receiving ICI, split by response to the treatment (n=11). **d-g)** Circular networks of the significant differences in magnitude (as calculated by Cohen’s D) of the relative contribution of immune marker expression in relevant cell types by cell-rich and cell-poor communities in responders (top) or non-responders (bottom) for *PDL1* and *PD1* (**d**), *CTLA4* and *CD86* (**e**), *LAG3* (**f**), and *TIM3* (**g**). Nodes that are labeled as “community” are colored light brown, while the other nodes are colored based on the cell types that are relevant for each immune marker. Edge colors denote proportion of expression differences (rich: steel blue; poor: magenta), and edge widths indicate the strength of associations based on Cohen’s D. All reported associations in panels c-g have been adjusted for using Benjamini-Hochberg (*p < 0.05). ns: p > 0.05 / *: p <0.05*).

Overall, the localization of cells expressing checkpoint genes (or their ligands) is more localized in specific tumour areas in responder patients, but it is more widespread -yet spatially organized- in non-responders (bottom), as evidenced by the higher number of edges in the latter (**Figure 7d-g**). In tumours from responders, the most significant association was the highest fraction of *PD1* and *CTLA4* expression coming from CD8 T cells in fibroblast-poor communities (**Figure 7d,e**), compared to CD8 T cells in fibroblast-rich communities. These findings contrast with those from tumours of non-responder patients, who showed many more connections for both *PDL1-PD1* and *CTLA4-CD86* pairs. In these tumours, communities rich in M1 macrophages, monocytes, or endothelial cells were the main contributors to *PDL1* and *PD1* expression (**Figure 7d**). Regarding *CTLA4* expression in T cells (CD4, CD8, and T regs), it was significantly higher in communities rich in monocytes and M1-macrophages, than in T cells located in communities poor in these two cell types (**Figure 7e**). Additionally, in the case of CD8 T cells, *CTLA4* expression is also biased towards epithelial-poor communities (**Figure 7e**), suggesting a potential intrinsic resistance mechanism. Finally, we explored the architecture of the expression of *LAG3* and *TIM3* (**Figure 7f,g**). The overarching architecture in responders is similar to that of the previous LR pairs, with selective expression in a few key communities in responders, and a wider, spatially structured, expression in non-responding patients. We identified a high expression of *LAG3* and *TIM3* in NK and CD8 T cells in fibroblast-poor communities of responders (**Figure 7f,g**). In contrast, in non-responders *LAG3* was mainly expressed in plasma cells with a strong bias towards M1 macrophage-rich communities, among others (**Figure 7f**). In these tumours, *TIM3* expression occurred in CD8 T cells, NK cells, or dendritic cells with a bias towards epithelial-poor, endothelial-, monocyte-, and M2-rich communities (**Figure 7g**).

These findings highlight the complex interplay between immune signaling and the unique cellular composition of different community types in responders, suggesting a highly localized regulation of immune activity that differs from that of non-responders and, therefore, may contribute to differential responses to ICI treatment.

## Discussion

The DUTRENEO trial is a unique prospective study designed to identify which patients with MIBC are more likely to benefit from ICIs compared to standard NAC. This innovative approach is based on the TIS gene signature, a robust biomarker suggested to be able to optimize patient selection for ICI treatment^15^.

The pCR rate in patients who received ICI was similar to that achieved in a recent study^11^ using the same drugs/schedule in 28 cisplatin-ineligible MIBC patients (37.5%) or that of another study^12^ using nivolumab (a different PD1 inhibitor) with tremelimumab in 24 patients that yielded a pCR rate of 46%. Therefore, compared to studies in an unselected patient population, the TIS as used in the DUTRENEO trial did not result in a significantly improved responsiveness to ICI. The response rate was similar to that of patients who received NAC, regardless of their immune infiltration. A retrospective analysis of the data shows that the TIS does associate with the outcome of ICI-treated patients: using a higher TIS threshold to enroll patients in the TIS-high arm would have led to a higher response rate (**Figure 1e**). At the TIS threshold applied (>6), 37% of patients achieved a pCR; we estimate that this rate would have increased to 42% or 50% had we considered cases with a TIS >7 or >8, respectively. However, using these thresholds, 50% (5/10) and 70% (7/10) of patients randomized to the ICI arm who achieved a pCR would not have had the opportunity to receive immunotherapy (**Figure 1e**).

Overall, our findings highlight the challenges of selecting patients to receive ICI based on a single biomarker, even when it has been shown to be robust in retrospective analyses and to associate with the desired outcome and biological hypotheses in a prospective trial. A few suggestions for future trials that aim to prospectively test patient stratification schemas include: i) using multiple thresholds in their design, ii) applying continuous rather than dichotomous variables to estimate response likelihood, iii) using combinations of biomarkers that are not highly correlated, and iv) combining biomarkers that are predictive of response (or lack thereof) to more than one type of therapy.

In this context, our multi-omic characterization of the DUTRENEO pre-treatment samples provides new insights into the biology of MIBC. We have identified new molecular features distinctly associated with response to neoadjuvant ICI or NAC. In our study, *PDL1* expression, assessed using IHC, the standard companion diagnostic biomarker, did not predict response to ICI; yet, there were significant differences in the expression of *PDL1* and *CTLA4* in CD4 T regulatory cells and dendritic cells in responders vs. non-responders. Additionally, beyond the cell type where checkpoints are expressed, we found that the composition of the cancer cell neighbourhoods was clearly different in ICI-treated patients who responded vs. those who did not. In tumours from the former, cancer cells were in close proximity to immune cells, particularly those from the adaptive arm. This observation is in line with other studies that have used spatial -omics platforms to characterize the relationships and distances between cancer and immune cells in patients who received immunotherapies in breast^27,28^, ovarian^29^, esophageal^30^ or lung cancer^31^, among others^32^. This suggests that this could, indeed, be another PanCancer biomarker, possibly in combination with bulk RNA signatures. Further validation of our findings, with integration with multiple biomarkers, is warranted.

Another potentially important result is that, among patients receiving chemotherapy, the pCR rate was higher for cold tumours than for hot tumours (53% vs. 30%, respectively). Cold tumours were enriched in the luminal consensus subtypes, but there is controversy on the association between molecular subtypes and response to platin-based NAC^33–36^. There is a need to identify robust biomarkers of response to NAC, and several ongoing trials are addressing this question. Our analyses showing that, in patients who did not respond to NAC, cancer cells explore a broader transcriptomic space in the absence of major genomic diversity (measured as CNV) points to phenotypic plasticity and phenotype-driven selection^37,38^ as a major driver of drug resistance in MIBC. Therefore, clones with multiple molecular consensus subtypes coexist in MIBC, as has been reported for other carcinomas^39,40^.

These observations highlight a central challenge moving forward: the biomarkers predictive of chemotherapy efficacy differ fundamentally from those associated with immunotherapy response. This divergence poses a significant hurdle when attempting to assess both sets of biomarkers concurrently in the same patient to guide treatment decisions. The complexity of this challenge is further amplified by the rapidly evolving therapeutic landscape in bladder cancer, which now includes emerging options such as anti-NECTIN4 antibodies and other antibody–drug conjugates.

Our study has some limitations, largely derived from being a phase 2 trial with a modest sample size. These include lack of stratification by clinical prognostic factors and prior BCG therapy and differences in the percentage of patients who completed the full pre-planned treatment doses. Even though this is the largest spatial transcriptomics report in MIBC until now, our results are based on the analysis of a relatively small number of samples.

In summary, while the TIS in its current application did not significantly enrich for ICI responders, our study provides critical insights into the spatial organization of the tumour microenvironment and the role of transcriptional plasticity in therapeutic resistance. By integrating multi-modal data and embracing a continuous, integrated approach to biomarker assessment, future research can pave the way for more precise patient stratification. Ultimately, developing models that reconcile these distinct biomarker profiles will be essential to determine which treatment modality—be it chemotherapy, immunotherapy, or targeted therapy—is most likely to benefit individual patients in an era of expanding therapeutic options.

## METHODS

### Study population, trial design, and statistical considerations

DUTRENEO is a multicenter, open-label, study to evaluate the antitumor activity of tremelimumab and durvalumab in comparison with standard platinum-based NAC in patients with predominant urothelial muscle-invasive bladder cancer defined by cT2-T4a and/or lymph node involvement (N1+) staging with residual disease after TURBT, no evidence of distant metastasis and eligible to receive a cisplatin-based treatment by Galsky’s criteria^41^. Additional inclusion criteria included Eastern Cooperative Oncology Group performance status of 0 or 1, absence of previous chemotherapy for muscle-invasive disease, adequate blood counts and liver function - including a calculated creatinine clearance of > 40 ml/min - and HbA1c ≤ 8%. Main exclusion criteria were histology of pure adenocarcinoma, pure squamous cell carcinoma, or predominant small cell carcinoma or sarcomatoid features and evidence of distant metastasis. Current or prior use of immunosuppressive medication for an immune-related disease was also considered an exclusion criterion.

Based on an estimated positivity rate of the TIS in half of the patients and to detect an improvement in pCR rate from 30% with standard cisplatin-based chemotherapy up to 50% with durvalumab + tremelimumab (α: 0.1; β: 0.1). It was considered that 22 patients in each arm would be needed in a first phase. If at least 8 pCR were observed among the first 22 patients recruited in the investigational ICI arm, 24 additional patients would be subsequently recruited in the ICI arm. A dropout rate of 10% was assumed, therefore the total sample size needed to fulfil the objectives of the trial were 99 patients. A prospective and centralized TIS based on 18-gene IFN- signaling-related expression (see Results) was obtained on TURBT samples to classify patients. One-hundred-one patients were screened for study eligibility. Of them, 28 resulted in screening failures [concurrent metastases (5); abnormal blood chemistries (2); abnormal QT interval (3); insufficient tumour sample to assess TIS (2); intercurrent infection (1); later refusal to participate (3); others (12, including 8 patients for which a slot was not available in the corresponding treatment arm)].

To establish a threshold to categorize tumors as TIS-high vs. TIS-low, we used TURBT samples (n=65) from a retrospective series of patients with MIBC who received cisplatin-based NAC in two hospitals participating in the DUTRENEO trial. The NanoString nCounter platform was used and TIS scoring was carried out following an established algorithm. A threshold that discriminated the samples with the lowest 30% scores was established to reach a balance between case enrichment and recruitment rate feasibility.

Subjects whose tumours were classified as cold were offered standard platinum-based NAC with gemcitabine 1250 mg/m^2^ on days 1 and 8, and cisplatin 70 mg/m^2^ on day 1, every 3 weeks for a total of three cycles or dose dense-MVAC with methotrexate 30 mg/m^2^ on day 1, vinblastine 3 mg/m^2^ on day 2, doxorubicin 30 mg/m^2^ on day 2, and cisplatin 70 mg/m^2^ on day 2 with associated granulocyte colony-stimulating factor (G-CSF) from day 3 to day 9, every 2 weeks for a total of four cycles, according to investigator’s choice. Patients whose tumours were classified as hot were randomized to receive either standard cisplatin-based NAC as mentioned above, according to investigator’s choice, or the combination of durvalumab (1500 mg) and tremelimumab (75 mg) intravenously every 4 weeks for a total of three cycles before surgery. A cystectomy was requested to be performed 4 to 8 weeks after the end of neoadjuvant treatment. The primary endpoint was pCR, defined as no evidence of residual disease based on independent pathological review of the surgical specimen of durvalumab plus tremelimumab arm in comparison with standard NAC after cystectomy in the intention-to-treat population. Secondary endpoints included disease-free survival, overall survival, and safety profile. Follow-up after surgery was performed following ESMO guidelines rules and site protocols. The sponsor of the study was the CRIS Cancer Foundation (https://criscancer.org/en/), thanks to an unrestricted independent grant from Astra Zeneca. The study was conducted in accordance with Good Clinical Practice Guidelines and the Declaration of Helsinki and in accordance with the applicable regulatory requirements, in particular the ICH tripartite harmonized standards for good clinical practice 1996 and the Royal Decree of Clinical Trials 1090/2015, by which clinical trials with medicinal products are regulated in Spain, which fully incorporates the Regulation (EU) No 536/2014 of the European Parliament and of the Council of 16 April 2014.

All patients signed the informed consent form to be enrolled in the trial, approved by the Ethics Committee of the Hospital Universitario Ramón y Cajal on September 03, 2018 (EudraCT number: 2017-002246-68).

Survival curves were derived using the Kaplan–Meier method and compared with log-rank tests. Progression-free survival and overall survival and were considered as outcomes. Hazard ratios and 95% confidence intervals were estimated with univariable and multivariable Cox PH models. Differences were considered statistically significant at two-sided p < 0.05 using non parametric statistical tests. All analyses were performed using R version 4.2.2 (https://www.r-statistics.com/tag/r-4-2-2/) unless other indicated.

### Immunohistochemical assays

Blocks from pre-treatment FFPE tumour samples were obtained from the participating hospitals. Sections (4µm) were deparaffinized and stained in a centralized laboratory with primary antibodies detecting PDL1 (rabbit monoclonal antibody E1L3N, Cell Signaling Technologies, #13684). Positive immunostaining scoring was performed blindly by an expert pathologist (AA), following standard protocols used for patient selection in the clinical setting^42^.

### Whole exome sequencing

H-E sections from pre-treatment FFPE TURBT samples were used to select areas with a high tumour cellularity (>60%). The same tumour regions were selected for macrodissection for WES and RNA-Seq. Data from tumour samples were analyzed from FASTQ files using Varca pipeline in tumoru-only mode (https://github.com/cnio-bu/varca). This pipeline implements GATK Best-practices workflow^43^. Briefly, reads were aligned to hg38 reference genome using BWA aligner and somatic mutations were called using Mutect2^44^. VCF files were filtered using a hg38 panel of normal samples for both exomes and whole genomes generated from the 1000 Genomes Project and annotated and transformed to Mutation Annotation Format (MAF) with vcf2maf^45^. Downstream analyses were carried out using R package maftools^46^. Due to the high degree of noise derived from FFPE tumour-only samples and the relatively small sample size in each group, we filtered the dataset using somatic mutations included in cBioportal^47^ (bladder cancer) to focus on the top mutated genes.

### Bulk RNA-Seq and bioinformatics analysis

RNA was extracted using the Covaris truXTRAC® FFPE total NA kit; RNA quality was assessed using bioanalyzer. Total RNA was converted into cDNA sequencing libraries with the “QuantSeq 3‘ mRNA-Seq Library Prep Kit (FWD) for Illumina” (Lexogen, Cat.No. 015). Briefly, library generation was initiated by reverse transcription with oligodT priming, followed by a random-primed second strand synthesis. A UMI Second Strand Synthesis module was used, whereby random primers featured a 6 nucleotide long Unique Molecular Identifier 5’ tag to be addressed at data analysis. Sequencing was performed using an Illumina NextSeq 550 instrument. Reads were aligned to GRCh38 genome using STAR aligner. UMIs were extracted and demultiplexed using UMI-tools software^48^ to improve transcript quantification and correct for PCR artifacts related to poor-quality samples. Transcript quantification was carried out with featureCounts and differential expression analysis was done using DEseq2 package^49^.

### Spatial Transcriptomics – Visium

The integrity of RNA extracted for bulk transcriptomics was assessed; the percentage of RNA fragments above 200 base pairs (DV200) was used to select samples. Briefly, tissue blocks were placed in the microtome (ThermoScientific HM340E) and trimmed to expose the tissue. Four 10 µm thick sections were placed in a chilled Eppendorf tube and the RNA was extracted using a protocol from Qiagen (Rneasy FFPE Kit 73504), following extraction, the product was analyzed by TapeStation. Samples with DV200 ≥ 30% were selected; 7 µm thick sections were placed in a water bath floating at 42 °C, collected and mounted onto a 6.5 × 6.5 mm capture area of the Visium Spatial Gene Expression slide (2000233, 10X Genomics). Capture areas contain approximately 5000 barcoded spots, providing an average resolution of 1–10 cells. After sectioning, the slides were dried at 42 °C for 3 h. The slides were then placed inside a slide mailer, sealed with parafilm, and left overnight at room temperature. Sections were deparaffinized by successive immersions in xylene and ethanol followed by H&E staining according to Demonstrated Protocol (CG000409, 10X Genomics).

Brightfield images were taken using a 10X lens on a Nikon Eclipse Ti2, images were stitched together using NIS-Elements software (Nikon) and exported as tiff files. After imaging, the glycerol and cover glass were carefully removed from the Visium slides by holding the slides in a water beaker and letting the glycerol diffuse until the cover glass detached and density changes were no longer visible in the water. The slides were then dried at 37 °C and incubated for decrosslinking for 60 min. Following this step, an overnight probe hybridization was performed, and libraries were prepared according to the Visium Spatial Gene Expression for FFPE User Guide (CG000407, 10X Genomics). Libraries were sequenced at Macrogen (Korea) using 1 lane of HiseqX 150PE (2x 150bp) per sample, applying 1% Phix. Sequencing was performed using the specific for FFPE following read protocol: read 1: 28 cycles; i7 index read: 10 cycles; i5 index read: 10 cycles; read 2: 50 cycles.

### Spatial Transcriptomics - Xenium in situ gene expression

The Xenium workflow began by sectioning 5 μm FFPE tissue sections onto the Xenium slides, according to the “Xenium FFPE Tissue Preparation Guide” (CG000578 Rev C, 10X Genomics). Briefly, the sections were cut, floated in an RNAse-free water bath at 42 °C, and carefully placed onto the capture area of a Xenium slide (PN-3000941). After sectioning, the slides were incubated at 42 °C for 3 h and kept in a sealed bag with desiccators at room temperature overnight.

The next day, the slides were incubated at 60 °C for 2 h, and were processed following the “Xenium In Situ for FFPE-Deparaffinization and Decrosslinking” protocol (CG000580 Rev D, 10X Genomics). After deparaffinization, the Xenium slides were assembled into Xenium cassettes, which allow for the incubation of slides on the Xenium Thermocycler Adapter inside a Thermocycler machine with a closed lid for optimal temperature control, and Decrosslinking step was performed.

The slides were then processed using the “Xenium In Situ Gene Expression” user guide (CG000582 Rev D, 10X Genomics). After decrosslinking PBS-T washes, the samples were incubated at 50 °C overnight for probe hybridization approximately 19 h with the Xenium Human Multi-tissue and Cancer Panel, which targets 377 human markers for bladder, breast, colon, heart, kidney, liver, lung, pancreas, etc. This was followed by a series of washing steps, a post-hybridization wash, a ligation step, an amplification step and finally the slides were treated with an autofluorescence quencher and the nuclei were stained.

At the end of the third day of sample preparation, the slides in the cassettes were loaded into the Xenium Analyzer. The first step in the instrument consists in a sample scan, where images of the fluorescent nuclei in each section are given, and these images allow the user to select and determine the regions to be included in the analysis.

We designed experiments to place between 3 and 4 samples per Xenium slide, with a total of 6 slides that correspond to 3 Xenium Runs in the Instrument, we were able to process and analyze a total of 20 samples.

Each run in the Xenium Analyzer lasted 4 days approximately. After that, the instrument was emptied of consumables and the slides carefully removed. PBS-T was added to the slides and a post-run H&E staining was performed.

### Data preprocessing

Gene expression matrices of all 20 Xenium samples generated by Xenium Analyzer (v1.5.0.3) were imported into Seurat (v5). Cells were filtered out if they had less than 4 genes in each sample, thus minimizing false positive downstream cell type assignment as a result of potentially imperfect cellular segmentation. Finally, the gene expression in each sample was normalized with the SCTransform function from Seurat.

As for Visium data, raw fastq files of all 33 samples were processed with the Space Ranger pipeline (version 1.3.1, 10x Genomics) and mapped to the pre-built human reference genome (GRCh38). The STutility R package was used to process the output of Space Ranger in a series of steps. First, genes with less than 5 UMIs were excluded. Second, mitochondrial and ribosomal genes, and non protein-coding genes were removed from the analysis. Genes that were kept were tagged as protein_coding, TR_V_gene, TR_D_gene, TR_J_gene, TR_C_gene, IG_LV_gene, IG_V_gene, IG_J_gene, IG_C_gene and IG_D_gene. Lastly, spots were filtered out if the number of features was lower than a threshold value established by the distribution in each sample. The gene expression of each filtered sample was then individually normalized by variance stabilizing transformation using Seurat’s SCTransform function.

### Cell type assignment (Xenium data)

Cells were annotated using the R package SingleR^50^, which performs unbiased identification of cell types in single cell transcriptomics data based on built-in reference transcriptomes of pure cell types. In our case, we used the “BlueprintEncodeData” reference dataset which included a number of different cell types. To minimize false positive assignments, we restricted the “main labels” of the reference to cell types that may populate bladder tumours: namely, epithelial cells, fibroblasts, smooth muscle, pericytes, neurons, monocytes, CD4 T cells, CD8 T cells, NK cells, B cells, macrophages, endothelial cells, dendritic cells, neutrophils, and eosinophils.

We ran the singleR function on the “counts” slot with the “fine labels” as the reference cell types. These included the aforementioned main labels as well as additional subtypes: namely, Tregs, CD4 Tcm, CD4 Tem, CD8 Tcm, CD8 Tem, plasma cells, naive B cells, memory B cells, class-switched memory B cells, macrophages M1, macrophages M2, and mv endothelial cells. We assessed the quality of assignment with the “plotScoreHeatmap” function from the SingleR package to inspect the confidence of cell clusters when assigned to a given (sub)type. Despite the high confidence, the limited number of genes in the Xenium panel led us to adopt a more robust approach for downstream analysis by grouping all CD4 subtypes (except for Tregs) under CD4 T cells and those of CD8 subtypes under CD8 T cells. Moreover, we grouped all B cell subtypes (except plasma cells) under B cells, and “mv endothelial cells” under “endothelial cells”. Similarly, pericytes and smooth muscles were joined together, whereas neutrophils and eosinophils were grouped under “granulocytes”. Accordingly, the final list of cell types which we could confidently represent in our Xenium samples are: epithelial cells, endothelial cells, fibroblasts, smooth muscle, neurons, granulocytes, monocytes, CD4 T cells, Tregs, CD8 T cells, NK cells, B cells, plasma cells, macrophages, macrophages M1, macrophages M2, and DCs. Cells that had a low confidence score by SingleR were labeled “unassigned”. Finally, we ran differential expression analysis for the final labels on the SCT-normalized, merged Xenium data of all 20 samples, using the FindAllMarkers with the default “Wilcoxon” test. The list provided complementary evidence for the confidence of the assigned cell types, where some of the classical or known markers could be clearly associated to the corresponding cell type.

### Cell-to-cell distance and neighborhood analysis

To calculate the median nearest-neighbour distance of each cell type to epithelial cells in each of the Xenium samples, we used the frNN function from the “dbscan” R package. We first established a 200-µm radius around each epithelial cell and calculated the distance of the nearest-neighbouring cell type to each epithelial cell within that radius. Then, we took the median nearest-neighbour distance of each cell type to all epithelial cells within a sample. As for the neighbourhood analysis, we counted the number of cells of any given type within a 25 µm radius of each cell (**Extended Data Figure S7c**), yielding a neighbourhood count matrix for each sample (**Figure 5a**). Next, we used MiniBatchKmeans function from the R package “ClusterR” (batch_size = 500, max_iters = 500) to get the total within-cluster sum of squares (wcss) to determine an optimum number of clusters for each sample’s neighborhood matrix using the elbow method (**Extended Data Figure S8**). We performed k-means clustering on the neighborhood matrix with the kmeans function in base R, based on the selected number of clusters “k” (i.e. communities) for each sample.

To establish if a community is “rich” (enriched), “neutral”, or “poor” (depleted) in any given cell type, we first created Seurat objects (specifying min.feature = 1) for each of the neighbourhoood count matrices and normalized them with SCTransform. Then, we set the clusters (communities) as the identity in these objects, and ran FindAllMarkers function from Seurat (test.use = “wilcox”, logfc.threshold = 0, min.pct = 0, and min.diff.pct = 0) on the normalized neighbourhood cell type assay. To systematically represent the enrichment status of each cell type within at least one community across all samples for downstream analysis, we adopted a lenient classification approach. Specifically, avg_log2FC values greater than, equal to, or less than zero were categorized as “rich”, “baseline”, or “poor”, respectively.

### Assigning the neoplastic and TME compartments (Visium)

The “ESTIMATE” R package was used to quantify the neoplastic cell purity within each spot in the Visium datasets. We successfully used this tool previously on a panCancer Visium dataset, with certified pathologists attesting to the accuracy of neoplastic purity assignment^25^. To perform the quantification, we extracted the gene expression of each spot and turned it into GCT format, and ran the “estimate” function that computed the stroma, immune, and ESTIMATE scores. We then ran k-means clustering on these ESTIMATE scores to create five clusters within each sample. The cluster with the lowest ESTIMATE values corresponded to the maximum purity of neoplastic cells, whereas that of the highest values corresponded to the minimum purity of neoplastic cells (i.e., TME). To test whether spots from cluster #3 could be further disambiguated by assigning them to either the cancer (clusters 1&2) or the TME (clusters 4&5) compartments, we ran differential gene expression between cancer and TME compartments using FindMarkers function from Seurat. Genes whose avg_log2FC >1 were considered up-regulated in the cancer compartment, whereas those with avg_log2FC <-1 were considered up-regulated in the TME compartment. Next, we calculated the average expression of all genes from each list separately in each spot of cluster #3, followed by taking the log2 of the division between the average expression of cancer genes and TME genes for each spot. Finally, if the log2FC value was >1 the spot was assigned to the cancer compartment, whereas if it was < −1 it was assigned to the TME compartment. Spots that had log2FC values between −1 and 1 were kept as cluster #3 (i.e., buffer).

### Consensus molecular classification of MIBC (Visium)

We used the consensusMIBC R package^20^ to assign class labels according to the consensus molecular classification of MIBC. Visium datasets were normalized with “LogNormalize’’ from Seurat as it was the recommended normalization method in the documentation of this tool. We then ran the getConsensusClass function in each sample on the normalized gene expression pseudo-bulk matrix created from the cancer compartment assigned by ESTIMATE. The same function and approach were used to identify the molecular classification of each clone reported by inferCNV.

### Quantifying copy number alterations (Visium)

The inferCNV method was used to detect copy number alterations (CNA) in each spot based on gene expression. Raw gene expression counts in the tumour compartments that we established based on ESTIMATE were used to infer CNA in the cancer compartment while the TME compartment served as a reference. The “Buffer” spots were excluded from this analysis. The inferCNV function was executed with the following parameters: run(object_infCNV, cutoff=0.1, cluster_by_groups=F, plot_steps=T, denoise=T, no_prelim_plot=F, k_obs_groups = 1, HMM=T, leiden_resolution = 0.005, BayesMaxPNormal = 0.2). The results were obtained from step 17, which included spot groups from the dendrogram and the state of each gene. Groups of spots with less than 10 were considered outliers and were therefore removed from downstream analysis. We calculated the genomic distance between each pair of groups within a sample by determining the number of genes with the same CNV state divided by the total number of genes used to infer CNV states (Jaccard Similarity Index score), followed by merging the groups that were found identical to be a single clone.

### Building and quantifying Cancer Hallmark gene signatures

The process of assigning genes to Cancer Hallmarks and quantifying their activity was previously described^25^. In brief, pathways from the Pathway Commons database were curated and filtered to exclude those that were too small, too large, or irrelevant, ensuring a refined list for analysis. A two-stage process was applied to assign pathways to Hallmarks. Initially, pathways were selected based on specific keywords associated with each Hallmark. This was followed by a refinement step that reduced overlap across Hallmarks to ensure specificity. The resulting assignments were manually reviewed to confirm their accuracy and relevance. Hallmark activity scores were calculated using Seurat’s AddModuleScore on normalized datasets, scaled, and centered within each sample.

### Graphical figures

Individual panels with statistical data were generated with the R packages “ggplot2”, “pheatmap”, “ComplexHeatmap”. Circular network plots were generated with the R packages “ggraph” and “igraph”. Figures 1a, 3h, and 5a-b were generated using BioRender, and all individual panels were put together using Inkscape.

### Statistical analyses

Description of the qualitative data was carried out with absolute frequencies and corresponding percentages. The quantitative data were studied by using the mean ± standard deviation, the median, the minimum and maximum values. Percentages of responses were estimated using 95% confidence intervals. Comparisons between patient subgroups were made via Fisher’s exact test for categorical variables and t test for continuous variables. All correlations were calculated using Spearman test.

## Data availability

All data is available upon reasonable request from medical or research professionals for specific purposes. WES and RNA-Seq data are being deposited in the Gene Expression Omnibus.

## Acknowledgments

We thank Leticia Rodriguez, Natalia del Pozo, and the CNIO Histopathology and Genomics Unit for excellent technical support; Jedd Wolchock and Antoni Ribas for discussions; Javier Carmona and Joaquin Mateo for valuable comments to the manuscript; and the CRIS Cancer Foundation and APICES for their contributions to the implementation of the study.

## Author contributions

Conception and design: EG, MS, EA, DC, AF, EPP, NM, ID, EPP, and FXR

Financial support: EG, DC, AF, EPP, NM, EPP, and FXR

Administrative support: FXR

Patient recruitment and provision of study materials: EG, OR, DC, JP, AF, TAG, MAM, JB, MAC, MD, IG, SGM, FG, PM, AP, and ID

Collection and assembly of data: EG, MS, DG, EA, AA, JMdV, MAS, PG, JFG, DG, MM, APr, EPP, NM, and FXR

Data analysis and interpretation: EG, MS, DG, EA, AA, JMdV, MAS, RB, AM, JFG, DG, MM, JMP, NM, ID, EPP, and FXR

Manuscript writing: EG, MS, EA, EPP, and FXR

Final approval of manuscript: EG, MS, DG, EA, OR, AA, DC, JP, JMdV, AF, TAG, MAS, RB, AM, MAM, JB, MAC, MD, PG, IG, JFG, XGM, DG, FG, MM, PM, JMP, APi, APr, NM, ID, EPP, and FXR

Accountable for all aspects of the work: EG, EP, and FXR

