## Supplementary Tables and Figures for "Spatial biomarkers of response to neoadjuvant therapy in muscle-invasive bladder cancer: the DUTRENEO trial"

**Index Extended Data**

Extended Data Table 1. Treatment-related adverse events

Extended Data Figure S1. Study design, genomic, and molecular landscape of tumours.

Extended Data Figure S2. Survival analysis and association of PDL1 protein expression with response.

Extended Data Figure S3. Pathway analysis of bulk RNA-Seq data.

Extended Data Figure S4. Cell type composition across treatments and TIS status (Xenium).

Extended Data Figure S5. Association of genomic and phenotypic intratumoral heterogeneity (ITH) with response (Visium).

Extended Data Figure S6. The relative share of cell types to checkpoint expression across the whole section of each ICI-treated tumour (Xenium).

Extended Data Figure S7. Association with response based on distances and cell type composition (Xenium).

Extended Data Figure S8. Elbow method for optimum number estimation of communities (k).

Extended Data Figure S9. Niche co-expression scheme and association with response.

**Supplementary Table 1. Treatment-related adverse events**

|  | **TIS-high (hot)**  **cICI**  **N=30** | | | | | | **TIS-high (hot)**  **NAC**  **N=22** | | | | | | **TIS-low (cold)**  **NAC**  **N=21** | | | | | |
| --- | --- | --- | --- | --- | --- | --- | --- | --- | --- | --- | --- | --- | --- | --- | --- | --- | --- | --- |
|  | **Grade 1-2** | | **Grade 3-4** | | **Total** | | **Grade 1-2** | | **Grade 3-4** | | **Total** | | **Grade 1-2** | | **Grade 3-4** | | **Total** | |
|  | **N** | **%** | **N** | **%** | **N** | **%** | **N** | **N** | **N** | **%** | **N** | **%** | **N** | **%** | **N** | **%** | **N** | **%** |
| **General disorders and administration site conditions** |  |  |  |  |  |  |  |  |  |  |  |  |  |  |  |  |  |  |
| Asthenia | 12 | 40.0 | 1 | 3.3 | 13 | 43.3 | 13 | 59.1 | 1 | 4.5 | 14 | 63.6 | 10 | 47.6 | 2 | 9.5 | 12 | 57.1 |
| Decreased appetite | 0 | 0.0 | 0 | 0.0 | 0 | 0.0 | 4 | 18.2 | 0 | 0.0 | 4 | 18.2 | 9 | 42.9 | 0 | 0.0 | 9 | 42.9 |
| Mucosal inflammation | 1 | 3.3 | 0 | 0.0 | 1 | 3.3 | 1 | 4.5 | 1 | 4.5 | 2 | 9.1 | 2 | 9.5 | 2 | 9.5 | 4 | 19.0 |
| **Gastrointestinal disorders** |  |  |  |  |  |  |  |  |  |  |  |  |  |  |  |  |  |  |
| Nausea | 1 | 3.3 | 0 | 0.0 | 1 | 3.3 | 13 | 59.1 | 0 | 0.0 | 13 | 59.1 | 9 | 42.9 | 0 | 0.0 | 9 | 42.9 |
| Vomiting | 0 | 0.0 | 0 | 0.0 | 0 | 0.0 | 4 | 18.2 | 0 | 0.0 | 4 | 18.2 | 5 | 23.8 | 0 | 0.0 | 5 | 23.8 |
| Constipation | 1 | 3.3 | 0 | 0.0 | 1 | 3.3 | 4 | 18.2 | 0 | 0.0 | 4 | 18.2 | 4 | 19.0 | 0 | 0.0 | 4 | 19.0 |
| Diarrhoea | 2 | 6.7 | 0 | 0.0 | 2 | 6.7 | 1 | 4.5 | 0 | 0.0 | 1 | 4.5 | 4 | 19.0 | 0 | 0.0 | 4 | 19.0 |
| Dyspepsia | 0 | 0.0 | 0 | 0.0 | 0 | 0.0 | 1 | 4.5 | 0 | 0.0 | 1 | 4.5 | 3 | 14.3 | 0 | 0.0 | 3 | 14.3 |
| **Blood and lymphatic system disorders** |  |  |  |  |  |  |  |  |  |  |  |  |  |  |  |  |  |  |
| Thrombocytopenia | 0 | 0.0 | 0 | 0.0 | 0 | 0.0 | 5 | 22.7 | 3 | 13.6 | 8 | 36.4 | 6 | 28.6 | 1 | 4.8 | 7 | 33.3 |
| Neutropenia | 0 | 0.0 | 0 | 0.0 | 0 | 0.0 | 1 | 4.5 | 5 | 22.7 | 6 | 27.3 | 5 | 23.8 | 2 | 9.5 | 7 | 33.3 |
| Anaemia | 0 | 0.0 | 0 | 0.0 | 0 | 0.0 | 4 | 18.2 | 0 | 0.0 | 4 | 18.2 | 8 | 38.1 | 1 | 4.8 | 9 | 42.9 |
| Platelet count decreased | 0 | 0.0 | 0 | 0.0 | 0 | 0.0 | 1 | 4.5 | 1 | 4.5 | 2 | 9.1 | 2 | 9.5 | 2 | 9.5 | 4 | 19.0 |
| Neutrophil count decreased | 0 | 0.0 | 0 | 0.0 | 0 | 0.0 | 0 | 0.0 | 2 | 9.1 | 2 | 9.1 | 1 | 4.8 | 3 | 14.3 | 4 | 19.0 |
| **Skin and subcutaneous tissue disorders** |  |  |  |  |  |  |  |  |  |  |  |  |  |  |  |  |  |  |
| Alopecia | 1 | 3.3 | 0 | 0.0 | 1 | 3.3 | 5 | 22.7 | 0 | 0.0 | 5 | 22.7 | 5 | 23.8 | 0 | 0.0 | 5 | 23.8 |
| Pruritus | 7 | 23.3 | 0 | 0.0 | 7 | 23.3 | 1 | 4.5 | 0 | 0.0 | 1 | 4.5 | 0 | 0.0 | 0 | 0.0 | 0 | 0.0 |
| Rash | 5 | 16.7 | 0 | 0.0 | 5 | 16.7 | 0 | 0.0 | 1 | 4.5 | 1 | 4.5 | 0 | 0.0 | 0 | 0.0 | 0 | 0.0 |
| Palmar-plantar erythrodysesthesia syndrome | 0 | 0.0 | 0 | 0.0 | 0 | 0.0 | 0 | 0.0 | 0 | 0.0 | 0 | 0.0 | 1 | 4.8 | 0 | 0.0 | 1 | 4.8 |
| **Nervous system disorders** |  |  |  |  |  |  |  |  |  |  |  |  |  |  |  |  |  |  |
| Dysgeusia | 0 | 0.0 | 0 | 0.0 | 0 | 0.0 | 2 | 9.1 | 0 | 0.0 | 2 | 9.1 | 8 | 38 | 0 | 0.0 | 8 | 38.1 |
| **Metabolism and nutrition disorders** |  |  |  |  |  |  |  |  |  |  |  |  |  |  |  |  |  |  |
| Hypomagnesaemia | 0 | 0.0 | 0 | 0.0 | 0 | 0.0 | 0 | 0.0 | 0 | 0.0 | 0 | 0.0 | 3 | 14.3 | 0 | 0.0 | 3 | 14.3 |
| **Endocrine disorders** |  |  |  |  |  |  |  |  |  |  |  |  |  |  |  |  |  |  |
| Hyperthyroidism | 4 | 13.3 | 0 | 0.0 | 4 | 13.3 | 0 | 0.0 | 0 | 0.0 | 0 | 0.0 | 0 | 0.0 | 0 | 0.0 | 0 | 0.0 |

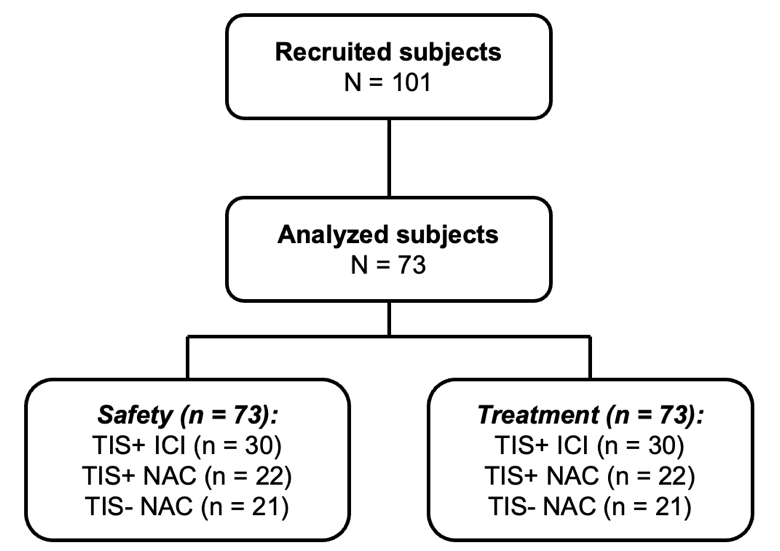

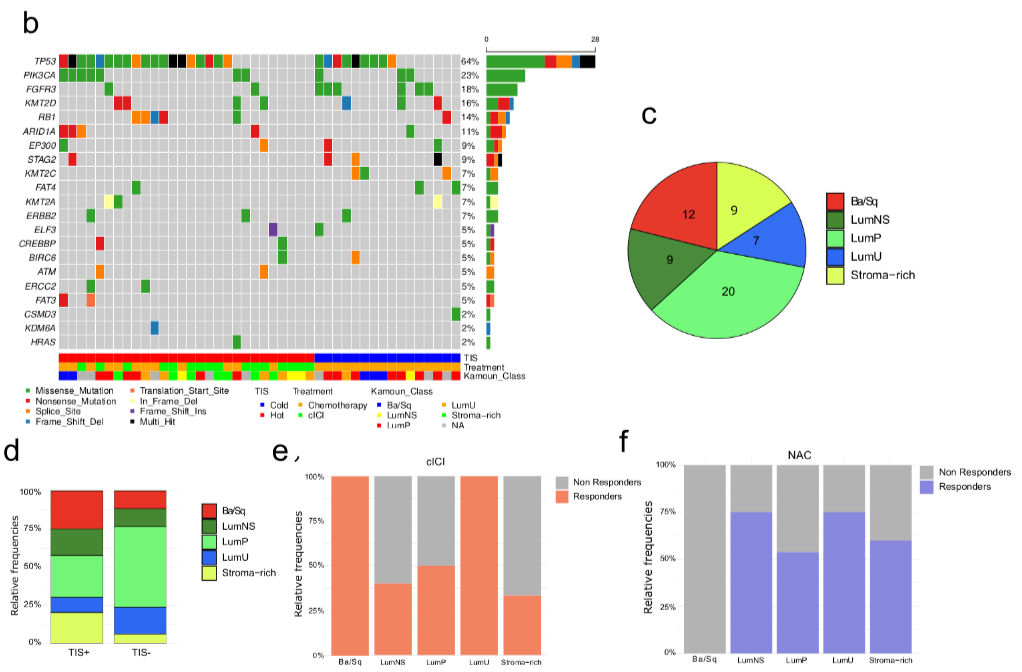

a

**Extended Data Figure S1. Study design, genomic, and molecular landscape of tumours. a,** DUTRENEO study design detailing the number of patients who underwent treatment administration and safety run-in. **b,** Frequency of cancer driver mutations with associated metadata on the TIS classification, treatment, and the consensus molecular subtype of each tumour. **c-f,** Tumour distribution according to the consensus molecular subtypes (**c**), split by TIS classification (**d**) or treatment received (**e,f**).

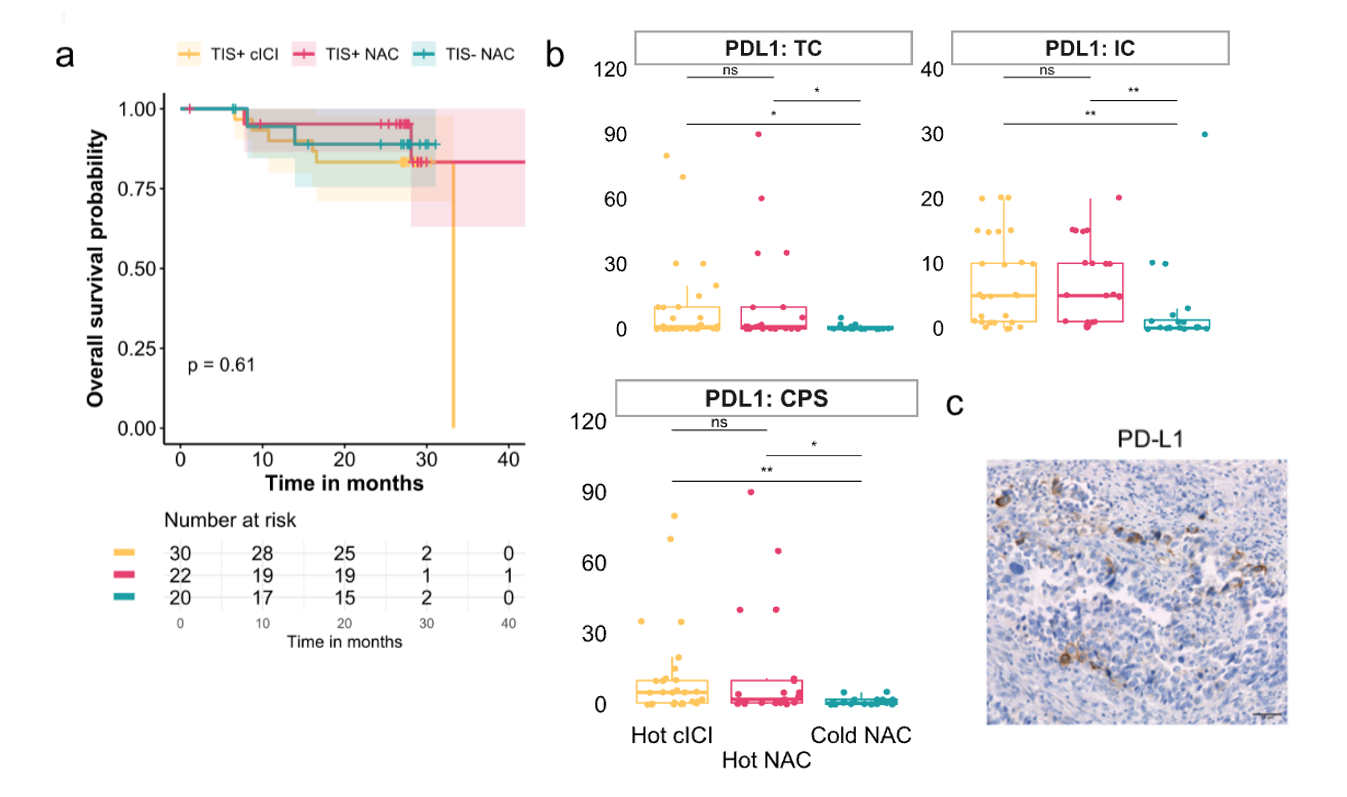

**Extended Data Figure S2. Survival analysis and association of PDL1 protein expression with the TIS. a,** Kaplan-Meier plot showing the probability of overall survival for patients included in the three treatment arms. **b,c,** Immunohistochemical (IHC) quantification of PDL1 protein levels in pre-treatment tumour samples: tumor cell score (TC, left), immune score (IC, right), as well as the combined score (CPS, bottom). **c**, representative IHC staining of PDL1 of one tumour section. *ns: p > 0.05, *: p <= 0.05, **: p <= 0.01, ***: p <= 0.001, ****: p <= 0.0001.*

**a**

**b**

**c**

**Extended Data Figure S3. Pathway analysis of bulk RNA-Seq data. a**, Pathways enriched among genes over-expressed (orange) or under-expressed (blue) in hot vs. cold tumours. **b, c,** Pathways enriched among genes over-expressed (orange) in tumours of responders vs. non-responders to ICI (**b**) and responders vs. non-responders to NAC (**c**).

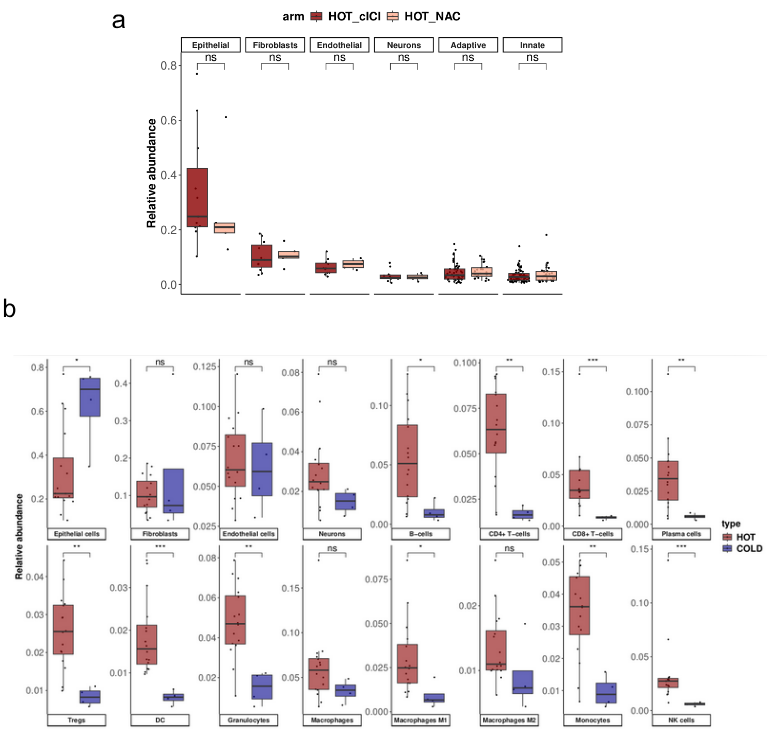

**Extended Data Figure S4. Cell type composition across treatments and TIS status (Xenium). a,** Relative cell type abundance in hot tumours from patients who received ICI vs. those who received NAC. Cell types belonging to the adaptive immune system are grouped together, as were those that belong to the innate system. **b,** Relative abundance of each individual cell type in hot vs. cold tumours. *ns: p > 0.05, *: p <= 0.05, **: p <= 0.01, ***: p <= 0.001, ****: p <= 0.0001.*

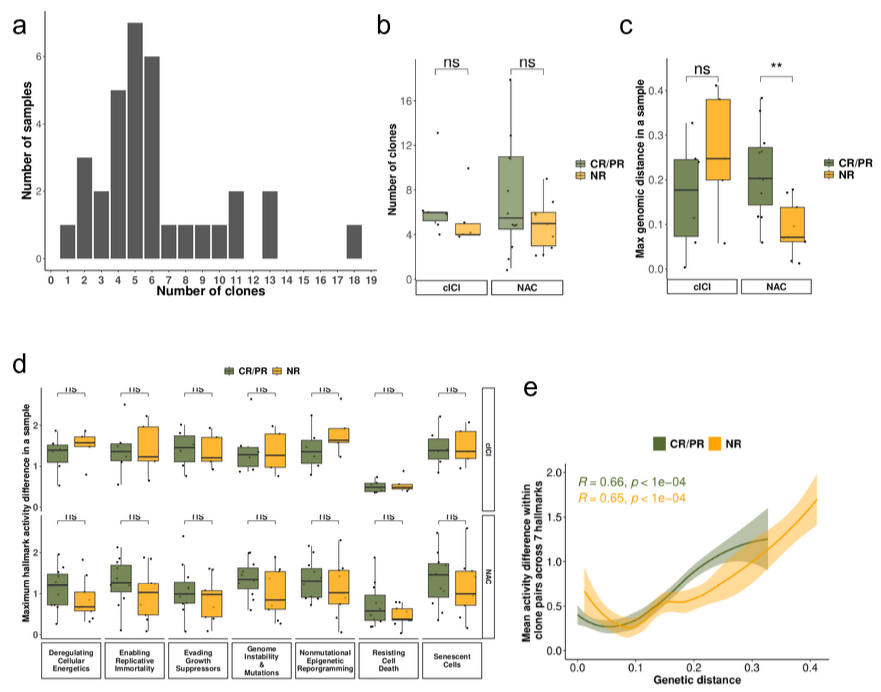

**Extended Data Figure S5. Association of genomic and phenotypic intratumoural heterogeneity (ITH) with response (Visium). a,b,** Number of CNV-based predicted clones for each sample (**a**) and lack of association with response in either ICI or NAC-treated patients (**b**). **c,** Maximum genetic distance between any two clones in a pair in each sample is associated with response to NAC but not ICI. **d,** Phenotypic ITH as measured by the maximum difference in the activity of each of the 7 neoplastic Hallmarks between between any two clones in a pair in each sample does not correlate with response to any of the two treatments. **e,** Scatter plot with a LOESS curve showing the interplay between genetic distance (1-JSI) and the mean difference in the activity between any two clones in a pair across the 7 neoplastic Hallmarks does not differ in responders vs. non-responders to ICI in the same way it does for NAC (NAC data shown in **Figure 3f**). *ns: p > 0.05, *: p <= 0.05, **: p <= 0.01, ***: p <= 0.001, ****: p <= 0.0001.*

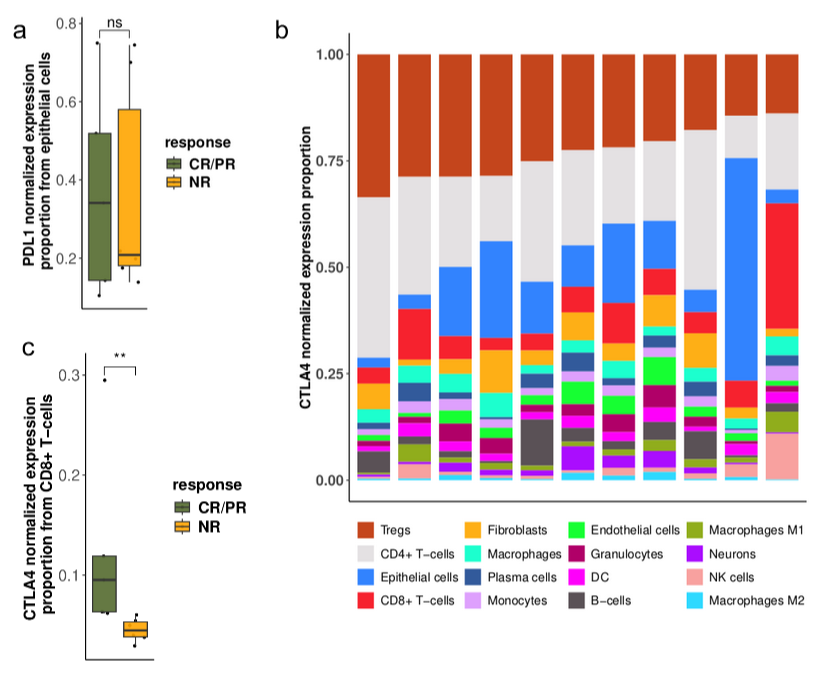

**Extended Data Figure S6. The relative contribution of cell types to checkpoint expression across the whole section of each ICI-treated tumour (Xenium). a,** Contribution of epithelial cells to the total expression of *PDL1* does not differ by response to ICI treatment. **b,** Contribution of cell types to the total expression of *CTLA4* shown for each ICI-treated patient. **c,** Contribution of CD8 T cells to the total expression of *CTLA4* is associated with response to ICI treatment. *ns: p > 0.05, *: p <= 0.05, **: p <= 0.01, ***: p <= 0.001, ****: p <= 0.0001.*

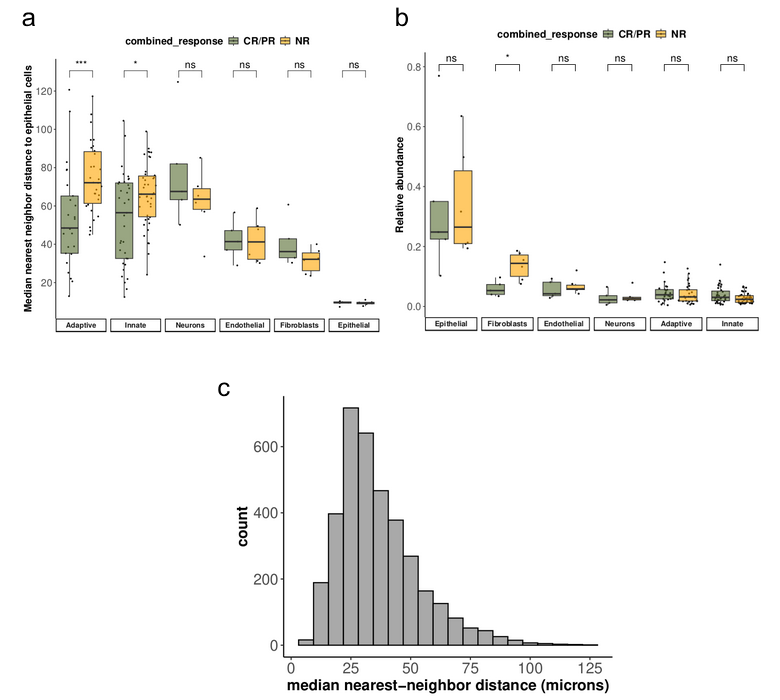

**Extended Data Figure S7. Association with response based on distances and cell type composition (Xenium). a,** Boxplot showing the median nearest neighbour distance of the different cell types to epithelial cells (calculated within a 200 μm radius) in all samples, split by response to ICI treatment. **b,** Relative cell type abundance split by response to ICI. Median distances of individual cell types pertaining to the adaptive and innate immune system were grouped under “adaptive” and “innate”, respectively. **c,** Histogram of the number of reported median nearest-neighbour distances of the cell types to epithelial cells in the previously established 200 µm radius from ICI-treated patients. *ns: p > 0.05, *: p <= 0.05, **: p <= 0.01, ***: p <= 0.001, ****: p <= 0.0001.*

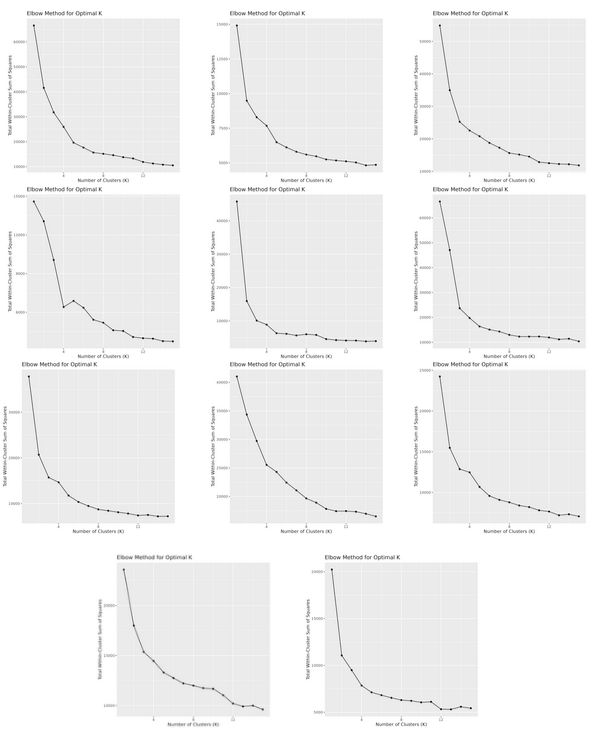

**Extended Data Figure S8. Elbow method for optimum number estimation of communities (k).** Elbow plots for each of the Xenium samples (n=11) were used to guide the selection of an optimum number of clusters to be used in k-means clustering of the neighborhood matrices. Selection was performed to ensure that the number selected would sufficiently high not to exclude important communities (> 4) and not excessively high (<9) to avoid noise.

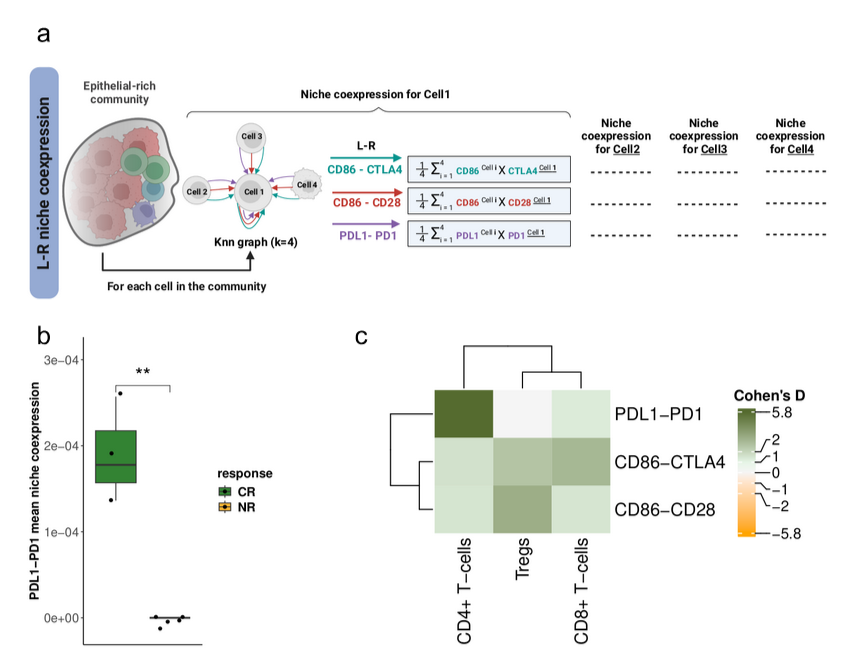

**Extended Data Figure S9. Niche co-expression scheme and association with response. a,** Using the “NICHES” method, the co-expression was calculated on each cell within epithelial-rich communities by taking the mean co-expression of a ligand and its receptor in the neighbouring three cells of a target cell. **b**, Mean niche co-expression of *PDL1-PD1* around CD4 T cells within epithelial-rich communities is significantly higher in complete responders (CR, n =3) compared to non-responders (NR, n =6) to ICI. **c,** Similar findings shown as a Cohen’s D heatmap (of CR vs NR) along with the mean niche co-expression of *CTLA4*-*CD86-* and *CD86-CD28* around the different T cells within epithelial-rich communities. *ns: p > 0.05, *: p <= 0.05, **: p <= 0.01, ***: p <= 0.001, ****: p <= 0.0001.*
